# Biallelic *BECN1* variants cause neurodevelopmental disorders via AMPA receptor dysregulation

**DOI:** 10.64898/2026.10.13.26365182

**Authors:** Kenta Kuramoto, Farid Ullah, Pingdong Liang, Idalis M. Ramirez, Lama AlAbdi, Bing Feng, Minami Sakurai, Myung-Su Kang, Huancheng Dong, Motunrayo J. Oyeyemi, Zuoheng Qin, Meghan A. Coghlan, Abhineet Sharma, Angelique Mercier, Anne McRae, Abdelhak Belmadani, Anis Contractor, Omar Abuyousef, Mais O. Hashem, Amal Alhashem, Sateesh Maddirevula, Fowzan S. Alkuraya, Yanlin He, Erica E. Davis, Congcong He

## Abstract

Genetic regulation of neurodevelopmental disorders that impair brain development and social interaction is complex and poorly understood. Although emerging evidence suggests a link between neurodevelopmental disorders and autophagy deficits, congenital autophagy gene disorders are rare in humans, and underlying molecular mechanisms remain unknown. Here we demonstrate that the autophagy protein BECN1 is a regulator of neuronal and social development in vertebrate species. We identify rare deleterious biallelic *BECN1* variants that cause neurodevelopmental disorders in two unrelated families. *Becn1* disruption also causes neuronal and social developmental defects in mouse and zebrafish models. BECN1 serves as an adaptor to recruit Arc to the E3 ubiquitin ligase UBE3A for polyubiquitination and degradation, thereby maintaining postsynaptic AMPA receptor (AMPAR) stability, AMPAR-mediated excitatory neurotransmission, and normal social interaction. Thus, our study illustrates new biology and a non-canonical mechanism by which BECN1 regulates neurodevelopment in humans, mice and zebrafish, via a crosstalk with the ubiquitin-proteasomal machinery.

## Introduction

Neurodevelopmental disorders are a group of congenital conditions hallmarked by abnormalities in brain development that involve impaired social interaction, communication, and intellectual function across the lifespan, such as autism spectrum disorders (ASD) (*1*). A link between Mendelian neurodevelopmental disorders and deficits in autophagy, a conserved lysosomal degradation pathway regulated by over forty autophagy-related (ATG) genes (*2*), has been reported recently for *ATG5* and *ATG7* (*3, 4*). However, congenital ATG gene disorders are rare and poorly understood in humans. The molecular mechanisms underlying the pathogenesis of these ATG gene diseases remain elusive, and treatment strategies are lacking.

*BECN1* (Beclin 1) is the mammalian ortholog of the yeast ATG gene *Atg6*/*Vps30*, and encodes a core regulator of autophagy and endocytic trafficking pathways (*2*). It is a component of the autophagy-inducing class III VPS34 phosphatidylinositol 3-kinase complex, which produces the lipid phosphatidylinositol (3)-phosphate for autophagosome formation. *BECN1* is generally known as a haploinsufficient tumor suppressor, a role established previously in mouse models (*5, 6*). Human *BECN1* is located on chromosome 17q21, a genomic locus that is often mono-allelically deleted in breast and ovarian cancers; however, because the genomic region proximal to *BECN1* contains additional tumor suppressors (*7*), it is challenging to exclusively determine the roles of BECN1 in human cancers. Inherited disease-causing variants in *BECN1* in humans are hitherto unreported. In addition, although ATG proteins are generally thought to participate in lysosome-mediated autophagy degradation, emerging evidence suggests that many of them also carry out diverse non-canonical functions beyond the degradation process (*8–12*). Yet, the non-canonical roles and mechanisms of ATG proteins, including BECN1, in neurons and neuronal regulation are poorly understood.

Here, we demonstrate with data from three vertebrate species that BECN1 is a regulator of neuronal and social development. Our findings reveal new biology and a non-canonical non-autophagic mechanism by which BECN1 regulates neurodevelopment in humans, mice and zebrafish, by stabilizing α-amino-3-hydroxy-5-methyl-4-isoxazolepropionic acid (AMPA) receptors (AMPARs) and AMPAR neurotransmission in hippocampal neurons, via a crosstalk with the ubiquitin-proteasomal machinery.

## Results

### Biallelic variants in *BECN1* cause neurodevelopmental disorders in two unrelated families

As an ongoing effort to identify a genetic diagnosis for families with neurodevelopmental conditions, two research centers independently identified two pedigrees with biallelic *BECN1* variants. Family 1 was comprised of recurrent pregnancies that both presented with normal prenatal ultrasound imaging at 20 weeks gestational age, but by 32-33 weeks showed severe bilateral ventriculomegaly, agenesis of the corpus callosum, and dysplastic cerebellum. Deliveries were at 36 and 35 weeks, respectively. The family 2 proband and her younger sister, both teenagers at the time of clinical assessment, had a history of global developmental delay, intellectual disability, abnormal communication and hypogenesis of the corpus callosum. We used clinical exome sequencing (ES) reanalysis as part of undiagnosed disorders research to identify rare biallelic variants in *BECN1* in these two unrelated families with overlapping neurodevelopmental symptoms (**Fig. 1A-B**, **Table 1**, **Supplementary Case Reports**). Both families harbor rare protein-impacting variants in *BECN1*; no other likely genetic causes for disease were identified. The affected female individuals in family 1 have compound heterozygous inherited *BECN1* variants: one missense variant (c.1123T>C; p.Cys375Arg, C375R), and one nonsense variant (c.976C>T; p.Gln326*, Q326*) that likely induces the loss of mRNA due to nonsense-mediated mRNA decay. The affected female individuals in family 2 harbor a homozygous *BECN1* missense variant (c.1115T>C; p.Phe372Ser, F372S). The biallelic variants identified in the patients are rare (minor allele frequencies: C375R, 2/1,401,492; Q326*, 0; F372S, 0) and are not present in homozygosity in public databases such as gnomAD (v4.1.0), bolstering the possibility for association with their clinical spectrum.

**Figure 1.**
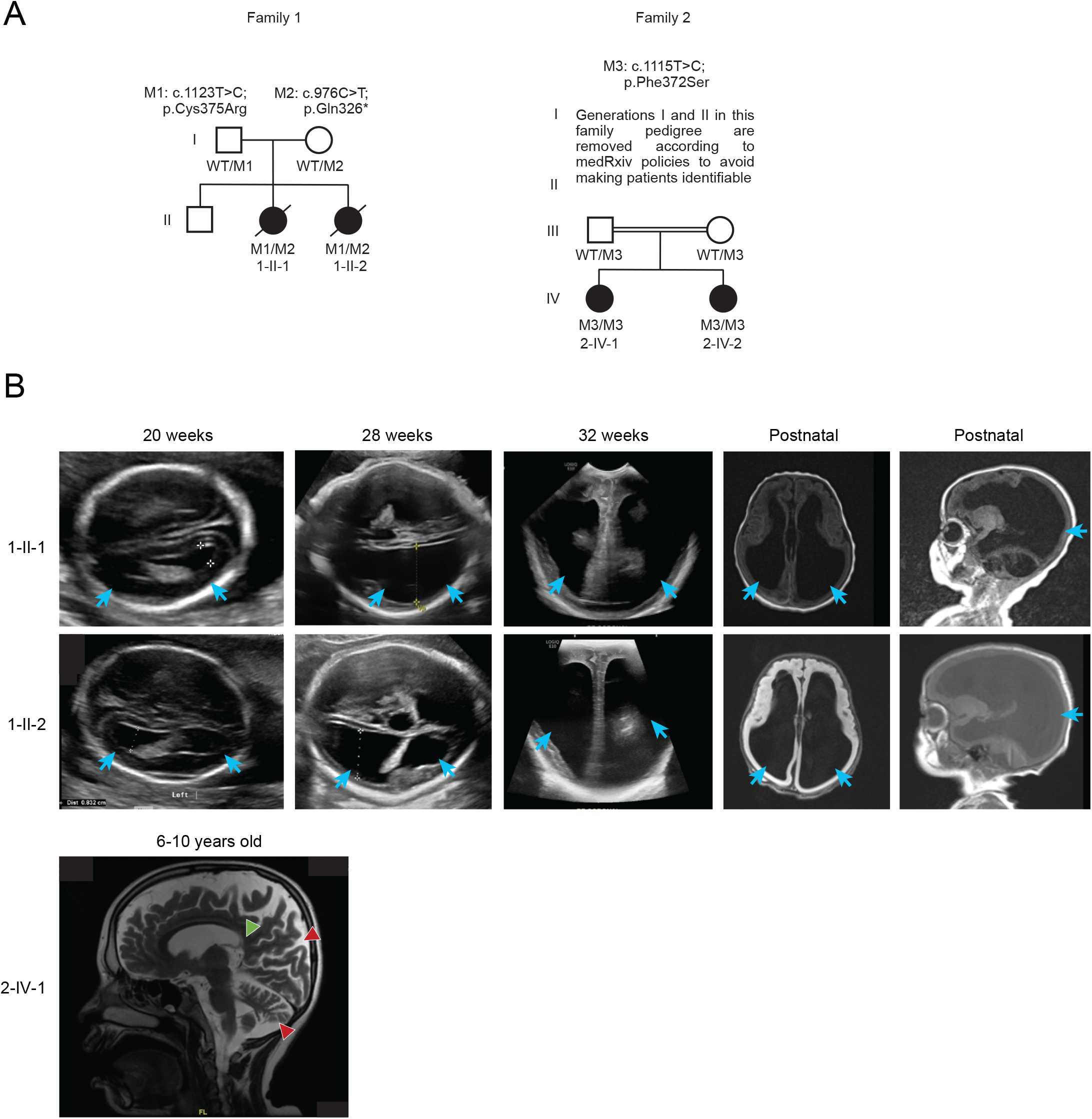
Identification of deleterious biallelic variants in *BECN1* causing neurodevelopmental disorders in two independent families. **(A)** Segregation of biallelic *BECN1* variants identified in two independent families; M1: c.1123T>C; p.Cys375Arg; M2: c.976C>T; p.Gln326*; M3: c.1115T>C; p.Phe372Ser. Filled shapes, affected individuals; unfilled shapes, healthy individuals; diagonal lines, deceased individuals; double horizontal lines, consanguineous union. **(B)** Representative prenatal brain ultrasound and postnatal brain MRI images of the affected individuals from family 1 and family 2. For family 1 images, blue arrows point to ventriculomegaly. For family 2 image, red arrowheads indicate diffuse supra and infratentorial brain atrophy noted with widening of cortical sulci and prominence of subarachnoid spaces and ventricular system; green arrowhead indicates diffuse thinning of corpus callosum noted more pronounced in the posterior body with hypogenesis of splenium of corpus callosum.

**Table 1.** Molecular and clinical features of affected individuals with biallelic variants in BECN1.

| Family # | Family 1 |  | Family 2 |  |
| --- | --- | --- | --- | --- |
| Individual ID # | 1-II-1 (AC0001-0100) | 1-II-2 (AC0001-0001) | 2-IV-1 (19DG0859) | 2-IV-2 (19DG0937) |
| Sex | female | female | female | female |
| Age at most recent examination | Removed to follow the medRxiv policy on patient identifying information |  |  |  |
| Ethnicity | Removed to follow the medRxiv policy on patient identifying information |  |  |  |
| Family history | negative | negative | first cousins/ familial | first cousins/ familial |
| Nucleotide alteration (NM_001313998.2; zygosity) | c.976C>T (Het-Mat); c.1123T>C (Het-Pat) | c.976C>T (Het-Mat); c.1123T>C (Het-Pat) | c.1115T>C (Hom-Mat/Pat) | c.1115T>C (Hom-Mat/Pat) |
| Protein alteration (NP_001300927.1) | p.Gln326*; p.Cys375Arg | p.Gln326*; p.Cys375Arg | p.Phe372Ser | p. Phe372Ser |
| Genomic coordinate (hg38) | chr17:42814528G>A; chr17:42811716A>G | chr17:42814528G>A; chr17:42811716A>G | chr17:42811724A>G | chr17:42811724A>G |
| Frequency in gnomADv4.1.0 | 0/ 1,614,012 alleles; 2/ 1,614,242 alleles; 0 Hom | 0/ 1,614,012 alleles; 2/ 1,614,242 alleles; 0 Hom | 0/ 1,614,012 alleles | 0/ 1,614,012 alleles |
| Domain | CCD; ECD-BARA | CCD; ECD-BARA | ECD-BARA | ECD-BARA |
| CADD Score | 42; 31 | 42; 31 | 32 | 32 |
| ACMG Classification | PM2, PP3, PS3<br>Likely Pathogenic.<br>PP3, PM2, PS3<br>Likely pathogenic | PM2, PP3, PS3<br>Likely Pathogenic.<br>PM2, PP3, PS3,<br>Likely pathogenic | PM2, PP3, PS3<br>Likely Pathogenic | PM2, PP3, PS3<br>Likely Pathogenic |
| Age at onset | Removed to follow the medRxiv policy on patient identifying information |  |  |  |
| Developmental milestones | Removed to follow the medRxiv policy on patient identifying information |  |  |  |
| Macrocephaly | Positive, 93%ile | Positive, 93%ile | Normal | Normal |
| Brain MRI | Agenesis of corpus callosum, dysplastic cerebellum, hypoplasia of vermis, enlarged ventricles | Agenesis of corpus callosum, dysplastic cerebellum, hypoplasia of vermis, enlarged ventricles | Hypogenesis of the corpus callosum with progressive cerebral volume loss | Hypogenesis of the splenium of corpus callosum.<br>No obvious other brain abnormality.<br>Incidental findings: enlarged adenoids and evidence of pan sinusitis as well as mastoiditis |
| Seizure(s) | Positive | Positive | Positive | No |
| Speech | N/A | N/A | Severe speech and language delay | Speech and language delay |
| Respiratory | Intubated due to poor respiratory effort | Progressive desaturation at 8 w of life | Normal | Normal |
| Cardiac | Negative | Atrial septal defect | No evaluation | No evaluation |
| Ocular | Bilateral optic disc atrophy | Blunted foveal reflex | Normal | No evaluation |
| Extremities | Bilateral postaxial buds on hands | Normal | Muscle stiffness | Muscle stiffness |
| Other | Negative | Negative | Congenital hypothyroidism and SLE | Congenital hypothyroidism |
| Other features | dolichocephaly | dolichocephaly | Multiple contractures and severe scoliosis to the left side | None |

**Abbreviations:** ACMG, American College of Medical Genetics Variant Interpretation (52); CADD, Combined Annotation-Dependent Depletion (53); CCD, coiled coil domain; ECD-BARA, Evolutionarily Conserved Domain / $\beta$ - α-repeated autophagy-specific; gnomAD, genome aggregation database accessed February 21, 2026; Het, heterozygous; Hom, homozygous; ID, identifier; m, months; Mat, Maternally inherited; MRI, magnetic resonance imaging; N/A, not applicable; Pat, Paternally inherited; PM2, Absent from controls (or at extremely low frequency if recessive) in Exome Sequencing Project, 1000 Genomes Project, or Exome Aggregation Consortium; PP3, Multiple lines of computational evidence support a deleterious effect on the gene or gene product (conservation, evolutionary, splicing impact, etc.); PS3, Well-established *in vitro* or functional studies supportive of a damaging effect on the gene or gene product; SLE, systemic lupus erythematosus; y, years.

### BECN1 is essential for neuronal and social development in zebrafish and mouse models

To reveal the roles of *becn1* in neurodevelopment, we used zebrafish models, which are a reliable proxy for modeling defects in neuroanatomy and head size observed in humans (*13, 14*). We performed reciprocal BLAST searches of *BECN1* against the zebrafish genome and identified a single ortholog with 79% identity and 88% similarity with human protein (**Fig. S1A**). To suppress zebrafish *becn1*, we generated two independent CRISPR gRNAs targeting *becn1* exons 2 and 7 and injected them with or without Cas9 protein into single-cell staged wild-type (WT) embryos. Similar to embryonic lethality induced by *Becn1* homozygous knockout (KO) in mice (*5, 6*), and a previously reported *becn1* zebrafish mutant (*15*), our stable germline homozygous *becn1* KO larvae (**Fig. S1A-E**) did not survive beyond 9-12 days post-fertilization (dpf), thus we characterized the neuroanatomy of F0 mosaic *becn1* mutants. By measuring area of the forebrain and region between the eyes using defined anatomical landmarks in F0 mosaic mutant animals, we found that head size is significantly increased in *becn1* mosaic animals generated by either gRNA (**Fig. 2A-B**, **Fig. S1F-G**), compared to non-injected control or gRNA-alone injected control. Wholemount acetylated tubulin staining further showed that compared to controls, *becn1* F0 mosaic animals have reduced optic tecta size, fewer intertectal axons and a significant decrease in cerebellum area, which are additional readouts for head size anomalies and corpus callosum hypoplasia (**Fig. 2C-D**, **Fig. S1H-I**).

**Figure 2.**
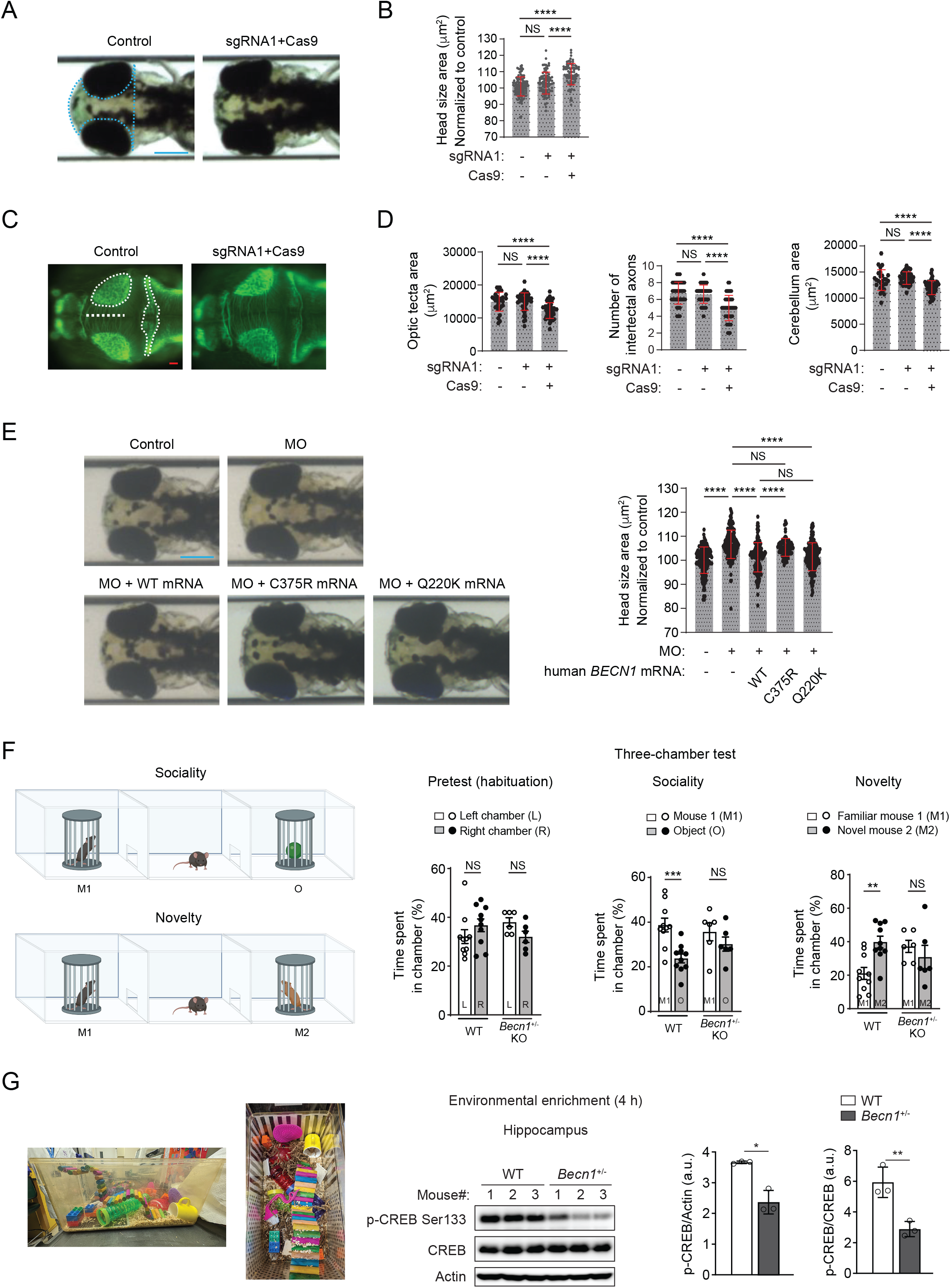
BECN1 is essential for neurodevelopment and social interaction in zebrafish and mouse models. **(A)** Representative dorsal bright field images of uninjected control and F0 mosaic mutant (sgRNA1) *becn1* zebrafish larvae at 3 dpf. Scale bar, 300 μm. **(B)** Quantification of dorsal head size area in uninjected controls and F0 mosaic mutants using landmarks shown with dotted blue line in panel A, n=70-95/condition. Similar results were produced in three biological replicates. **(C)** Representative dorsal fluorescent images of wholemount larvae stained with acetylated tubulin antibody for control or F0 mosaic animals targeted with sgRNA1+Cas9. Scale bar, 50 μm. **(D)** Quantification of optic tectum size (white circle), cerebellum area (white elongated dotted area), and number of intertectal axons (crossing the white dotted line) in panel C, n=50-70/condition. **(E)** Representative dorsal bright field images and quantification of the head size area of controls or larvae injected with *becn1* e5i5 morpholino (MO) with or without human WT *BECN1*, *BECN1*^C375R^, or *BECN1*^Q220K^ mRNA. Q220K is a negative control variant (rs138937709; 24 homozygotes in ∼1,614,172 individuals in gnomAD v4.1.0); n=70-107/condition. **(F)** *Becn1* heterozygous (+/-) (*Becn1*^+/-^) KO mice exhibit impaired social interaction by three-chamber tests. There was no preference for left (L) or right (R) side of the three-chamber unit in the habituation period for *Becn1*^+/-^ KO mice or WT littermates. Sociality was analyzed by time spent investigating mouse 1 (M1) vs. object (O). Preference for social novelty was analyzed by time spent investigating familiar mouse (M1) vs. novel mouse (M2). N=6-10. Two-way ANOVA with Bonferroni post-hoc test. (G) BECN1 is essential for hippocampal CREB phosphorylation (p-CREB, an indicator of synaptic excitatory activity) by environmental stimulation. (Left) Illustration of enviromental enrichment setup. (Right) Western blot (WB) analysis and quantification of p-CREB levels in the hippocampus of WT and *Becn1*^+/-^ KO mice after 4 hr of environmental enrichment. N=3. T-test. *, P<0.05; **, P<0.01; ***, P<0.001; ****, P<0.0001; NS, not significant. All error bars represent mean ± SEM.

Next, we validated F0 *becn1* mutant phenotype data with transient suppression models and tested missense variant pathogenicity with in vivo complementation. Using two independent splice-blocking morpholinos (MOs) targeting the splice donor sites of exon 5 and 6 (MO e5i5 and MO e6i6, **Fig. S2A-C**), we found that MO-mediated transient ablation of *becn1* also results in increased head size in zebrafish embryos in a dose-dependent manner (**Fig. S2D-E**). Co-injection of human *BECN1* WT mRNA or mRNA encoding a negative control benign population variant common in gnomAD v4.1.0 (c.658G>A; p.Gln220Lys, Q220K; human allele frequency: 6,080/1,614,172; 24 homozygotes), but not mRNA encoding the C375R missense variant, rescued head size in *becn1* morphants in a dose-dependent manner (**Fig. 2E**, **Fig. S2F**). In addition, acetylated tubulin staining showed that *becn1* morphants have reduced optic tecta size, fewer intertectal axons and a significant decrease in cerebellum area, which can be rescued when MO is co-injected with *BECN1* WT mRNA (**Fig. S2G-H**). Together, these results suggest that *becn1* is indispensable for normal brain development in zebrafish, and that the family 1 missense variant is pathogenic.

To further functionally study the role of BECN1 in neurodevelopment, we used mouse models to investigate neural development underpinning social interaction. Homozygous deletion of *Becn1* in mice causes embryonic lethality (*5, 6, 16, 17*), so we used the global *Becn1*^+/-^(heterozygous) mice. Using three-chamber tests, we found that *Becn1*^+/-^ KO mice of 12 weeks of age show defective social interaction in both sociality (preference for mouse over object) and novelty (preference for novel over familiar mouse) (**Fig. 2F**). Not all behavioral or psychiatric traits are affected in *Becn1*^+/-^ KO mice, as we detected no differences between *Becn1*^+/-^ KO and WT mice in striatum-mediated drug reward by cocaine conditioned place preference (**Fig. S3A**), spontaneous activity and exploratory behavior by open field (**Fig. S3B**), or anxiety by open field and elevated zero maze (**Fig. S3C**). *Becn1*^+/-^ KO mice also have comparable hippocampal neural spine morphology and density as WT mice (**Fig. S3D**). These data suggest that global haploinsufficiency of *Becn1* impairs social behavioral development. Given environmental and social enrichment increases neuronal activity and plasticity in the hippocampus (*18–20*), we analyzed neuronal stimulation via phosphorylation of the transcription factor CREB (cAMP-element binding protein), an indicator of synaptic excitatory activity. *Becn1*^+/-^ KO mice show reduced hippocampal p-CREB compared to WT littermates (**Fig. S3E**), and such reduction is more apparent after mice were subject to neuronal stimulation such as environmental enrichment (**Fig. 2G**, **Fig. S3E**), suggesting that BECN1 depletion impairs hippocampal neuronal stimulation. Together, these data demonstrate that BECN1 is a new critical regulator of neuronal and social activity and development in animal models.

### BECN1 is essential for postsynaptic AMPAR stability, AMPAR-dependent excitatory neurotransmission, and social interaction in hippocampal CA3 neurons

To further investigate the function of BECN1 in excitatory neurons, we generated tamoxifen-inducible CamKIIɑ-neuron *Becn1* conditional KO (CamKII^Becn1^-KO) mice. The CamKIIɑ promoter drives Cre recombinase expression robustly in cortex and hippocampal excitatory neurons, including CA1-CA3 pyramidal neurons (*21*). We confirmed markedly reduced BECN1 expression in the hippocampus and prefrontal cortex, but not the cerebellum or liver, of CamKII^Becn1^-KO mice after tamoxifen injection (**Fig. S4A**). CamKII^Becn1^-KO mice exhibited defects in sociality in three-chamber tests as global *Becn1*^+/-^ KO mice (**Fig. S4B**), further supporting that excitatory neuronal BECN1 is important for normal social development. Thus, we utilized electrophysiology approaches in these mice to determine the mechnanisms by which BECN1 regulates neurodevelopment and social interaction. AMPAR and NMDAR (N-methyl-D-aspartate receptors) are two major glutamate ionotropic receptors (ligand-gated ion channels) mediating excitatory transmission between neurons, where AMPAR mediates fast transmission (*22–24*) and NMDAR contributes to slower, longer-lasting transmission (*25, 26*). The cell-surface density of AMPAR and NMDAR on the postsynaptic membrane determines the onset and magnitude of excitatory neurotransmission. We recorded both AMPAR- and NMDAR-dependent miniature excitatory postsynaptic current (mEPSC) in hippocampal (CA1-CA3) neurons of WT control mice, global *Becn1*^+/-^ KO mice, and CamKII^Becn1^-KO mice (**Fig. 3A**). We found that the current amplitude of AMPAR mEPSC, but not NMDAR mEPSC, is significantly reduced in both global *Becn1*^+/-^ KO mice and CamKII^Becn1^-KO mice, while the frequencies (which are regulated presynaptically) remained unaltered (**Fig. 3B-C**), suggesting that *Becn1* KO disrupts postsynaptic AMPARs, but not NMDARs or presynaptic regulators. Such reduction in AMPAR mEPSC occurs most apparently in the hippocampal CA3 neurons, and less so in CA1 neurons, but not in CA2 neurons (**Fig. 3B-C**). Thus, these data suggest that BECN1 is required for AMPAR-mediated synaptic transmission in hippocampal CA3 neurons. To further test this hypothesis, we generated CA3 neuron-specific *Becn1* KO mice using two different methods, a genetic KO model (CA3^Becn1^-KO mice) by crossing *Becn1*^flox/flox^ mice with CA3-Cre (Grik4-Cre/G32-4 Cre) mice (**Fig. S5A**), and an AAV-mediated KO model (CA3^Becn1^-AAV mice) by bilaterally microinjecting control AAV (AAV2/9-CamKII-GFP) or Cre-AAV (AAV2/9-CamKII-GFP-Cre) into the hippocampal CA3 regions of *Becn1*^flox/flox^ mice (**Fig. S5B**). We found that compared to control mice, both CA3^Becn1^-KO mice and CA3^Becn1^-AAV mice showed impaired sociality and social novelty in three-chamber tests (**Fig. 3D**, **Fig. S5C**), supporting that BECN1 functions in CA3 neurons to regulate social interaction. In addition, similar to neural excitation in *Becn1*^+/-^ KO mice and CamKII^Becn1^-KO mice, Becn1 KO specifically in CA3 neurons decreases the amplitude of AMPAR-mEPSC, but not the amplitude of NMDAR-mEPSC or the frequencies of either AMPAR- or NMDAR-mEPSCs that are presynaptically regulated (**Fig. 3E-F**, **Fig. S5D-E**), further supporting that BECN1 in CA3 neurons is essential for postsynaptic AMPAR-mediated excitatory neurotransmission.

**Figure 3.**
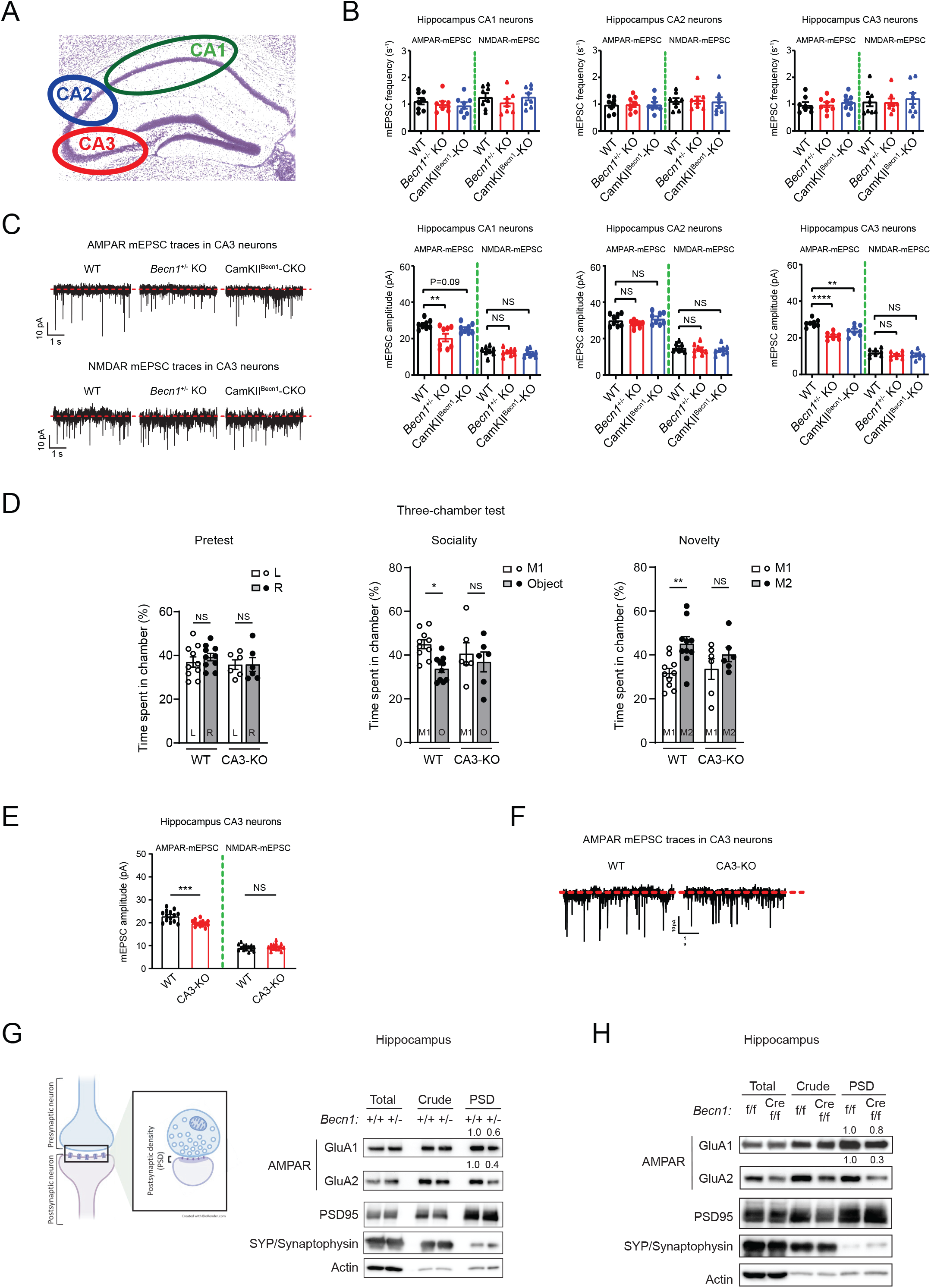
BECN1 regulates postsynaptic AMPAR stability, AMPAR-mediated excitatory neurotransmission, and social interaction in hippocampal CA3 neurons. **(A)** Illustration of hippocampal CA1, CA2 and CA3 neurons. **(B)** KO of *Becn1* impairs AMPAR mEPSC, but not NMDAR mEPSC, in hippocampal CA3 neurons. Measurement of the frequency and the amplitude of AMPAR-dependent mEPSC and NMDAR-dependent mEPSC in hippocampus CA1-CA3 neurons of WT mice, global *Becn1*^+/-^ KO mice, and conditional CamKII^Becn1^-KO mice. Two-way ANOVA. N=10-12 from 4 different mice in each group. **(C)** Representative traces of AMPAR mEPSC and NMDAR mEPSC in hippocampal CA3 neurons of WT mice, *Becn1*^+/-^ KO mice, and CamKII^Becn1^-KO mice. **(D)** Three-chamber test of WT control (*Becn1*^flox/flox^) and CA3 neuron-specific *Becn1* KO (CA3^Becn1^-KO mice) mice. N=6-10. Two-way ANOVA with Bonfferoni test. **(E)** Measurement of current amplitude of AMPAR-dependent mEPSC and NMDAR-dependent mEPSC in hippocampal CA3 neurons of WT and CA3^Becn1^-KO mice. **(F)** Representative traces of AMPAR mEPSC in hippocampal CA3 neurons of WT and CA3^Becn1^-KO mice. **(G)** Global BECN1 depletion reduces the level of AMPAR subunits GluA1 and GluA2 on the postsynaptic membrane of hippocampus. Synaptosomes and the postsynaptic density (PSD) were isolated from hippocampus pooled from 4 *Becn1*^+/-^ (+/-) mice and 4 WT littermates (+/+). **(H)** CamKIIɑ-Cre/ERT2-driven neuronal *Becn1* KO decreases AMPAR subunits on the postsynaptic membrane of hippocampus. Synaptosomes and PSD were isolated from hippocampus pooled from 4 CamKII^Becn1^-KO (Cre f/f) mice and 4 control (f/f) mice (tamoxifen-injected *Becn1*^flox/flox^ mice). Total, total lysates; Crude, crude synaptosomes; SYP/Synaptophysin (clone SVP38), presynaptic marker; PSD95, postsynaptic marker. Quantification against actin is shown above each protein band. *, P<0.05; **, P<0.01; ***, P<0.001; NS, not significant. All error bars represent mean ± SEM.

Next, to determine how BECN1 regulates postsynaptic AMPARs, we performed synaptosome studies (**Fig. S6A**). AMPARs are tetrameric complexes consisting of 4 subunits, GluA1-GluA4, among which GluA1 and GluA2 are the most abundant subunits in the hippocampus (*27, 28*). The GluA2 subunit is especially important, as it determines AMPAR Ca^2+^ permeability via the post-transcriptional glutamine (Q) to arginine (R) editing, which produces functional Ca^2+^-impermeable AMPARs. GluA1 and GluA2 mRNA transcription is unaltered in global *Becn1*^+/-^ KO mice or CamKII^Becn1^-KO mice (**Fig. S6B**). We validated the purification method for whole synaptosomes, presynaptic membranes, and postsynaptic membranes using SYP/Synaptophysin as a presynaptic marker, and PSD95 as a postsynaptic marker. We found that both AMPAR subunits, GluA1 and GluA2, show reduced levels on hippocampal postsynaptic membranes in both global *Becn1*^+/-^ KO mice (**Fig. 3G**) and CamKII^Becn1^-KO mice (**Fig. 3H**). In contrast, there are no detectable changes in the hippocampal synaptic levels of NMDARs in either global *Becn1*^+/-^ KO mice (**Fig. S6C**) or CamKII^Becn1^-KO mice (**Fig. S6D**) compared to control mice. These data suggest that BECN1 plays an important role in CA3 neurons to maintain the stability of AMPARs, but not other glutamate receptors, on postsynaptic membranes. Overall, we conclude that BECN1 functions in CA3 neurons to enhance postsynaptic AMPAR stability, AMPAR excitatory neurotransmission, and social development.

### BECN1 missense variants maintain protein structure and binding to the autophagy machinery, and do not detectably impact autophagy

BECN1 is conserved across species, and the two missense BECN1 variants identified in this study, F372S and C375R, reside in the evolutionarily conserved BARA (β-α-repeated autophagy-specific) domain at the C terminus of BECN1 (**Fig. S7A**). Via AlphaFold3 prediction, we did not detect evident abnormalities in protein folding or structure for either BECN1 mutant (**Fig. S7B**). In co-immunoprecipitation (co-IP) assays, BECN1^F372S^ and BECN1^C375R^ also maintained their interactions with known BECN1 binding partners important for autophagy and endocytic functions (*2*), including AMBRA1, UVRAG and VPS34 (**Fig. S7C**). The BECN1 inhibitor BCL2 binds and sequesters BECN1, thereby suppressing autophagy (*2*). Yet, we found that the interaction of BCL2 with BECN1^F372S^ and BECN1^C375R^ is modestly decreased, rather than enhanced (**Fig. S7D**), indicating that these BECN1 variants do not cause enhanced autophagy inhibition by BCL2. This is supported by normal autophagy flux in cells expressing BECN1^F372S^ or BECN1^C375R^, analyzed by degradation of the autophagy substrate p62 (the difference in p62 levels with and without lysosomal inhibition) (**Fig. S7E**). Thus, these data suggest that the BECN1 missense variants identified in affected humans likely maintain BECN1 protein structure, binding to the autophagy and endocytic machinery, and autophagy activity, suggesting a non-canonical mechanism of BECN1 in the regulation of AMPARs and social behavior.

### Identification of E3 ligase UBE3A and its substrate Arc as binding partners of BECN1

Because our data suggest that BECN1 enhances, rather than reduces, AMPAR levels and neurotransmission, and that human BECN1 variants do not detectably impact autophagy activity, we reasoned that autophagy disruption is not likely to be the predominant cause of the neurodevelopmental diseases in patient families, and that BECN1 regulates AMPARs via a non-canonical mechanism. Thus, to understand the pathogenic mechanism of BECN1 variants, we performed IP-mass spectrometry (IP-MS) analyses for BECN1-binding proteins. The study revealed diverse E3 ubiquitin ligases in the ubiquitination pathway as candidate binding partners of BECN1 or BECN2, or both (**Fig. S8A**). Among these candidates, we biochemically validated that several E3s, including UBE3A (Ubiquitin-protein ligase E3A), can be pulled down by BECN1 (**Fig. S8B**), and vice versa (**Fig. S8C**). *UBE3A* is the risk gene in Angelman syndrome, a neurodevelopmental disorder considered as a syndromic form of ASD (*29–32*). Although proteomics suggested that UBE3A can bind both BECN1 and BECN2, biochemical studies showed that endogenous UBE3A is a BECN1-binding, but not BECN2-binding, E3 (**Fig. 4A**). We also detected the interaction between endogenous UBE3A and BECN1 in vivo in the mouse hippocampus (**Fig. 4B**). These data suggest that the E3 UBE3A is a new BECN1 interaction partner in vitro and in vivo.

**Figure 4.**
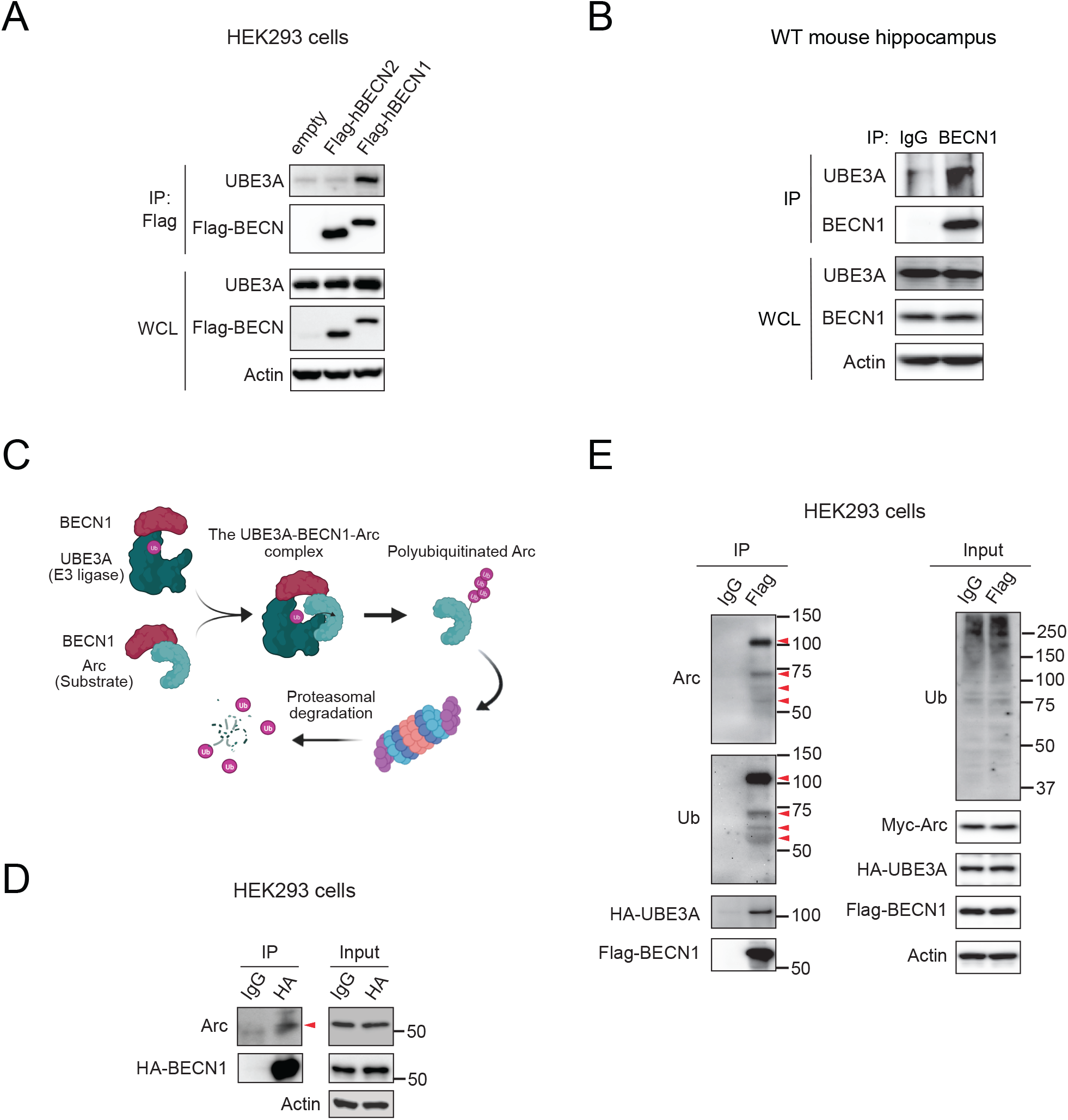
BECN1 is a novel binding partner of E3 UBE3A and its substrate Arc. **(A)** Endogenous UBE3A is co-immunoprecipitated with Flag-BECN1, but not Flag-BECN2, in HEK293 cells. **(B)** Endogenous UBE3A is co-immunoprecipitated with endogenous BECN1 in WT mouse hippocampus. WCL, whole cell lysates. **(C)** Proposed model of the mechanism by which BECN1 regulates Arc protein levels. BECN1 interacts with both the E3 ligase UBE3A and the substrate Arc, serving as an adaptor to facilitate UBE3A-mediated Arc polyubiquitination and proteasomal degradation. **(D)** BECN1 pulls down Arc in HEK293 cells transfected with HA-BECN1 and Myc-Arc, via co-IP. **(E)** BECN1 interacts with both unmodified and polyubiquitnated Arc, as well as UBE3A. HEK293 cells transfected with Myc-Arc, HA-UBE3A and Flag-BECN1 were treated with the proteasomal inhibitor MG132 (10 μM) prior to co-IP. Red arrowheads indicate Arc and polyubiquitinated Arc. Ub, ubiquitin.

We reasoned that there are two possible mechanisms for BECN1-UBE3A interaction: BECN1 either binds UBE3A and regulates its overall E3 activity, or recruits a specific substrate to UBE3A for ubiquitination by interacting with both the E3 and the substrate (**Fig. 4C**). Although the exact role and mechanism of UBE3A have yet to be fully determined, one reported UBE3A ubiquitination substrate is Arc/Arg.3.1 (Activity-regulated cytoskeleton-associated protein) (*33, 34*). However, a canonical E3 ligase-substrate interaction between UBE3A and Arc is not detected in either the cortex or the hippocampus (*35*), suggesting that new additional regulators may participate in UBE3A-mediated Arc ubiquitination. By co-IP, we discovered that Arc/Arg3.1 interacts with BECN1 in cells (**Fig. 4D**). Importantly, we found that BECN1 pulls down both unmodified (∼55 kDa) and polyubiquitinated (higher molecular-weight bands) Arc, together with the E3 UBE3A (**Fig. 4E**), suggesting that BECN1 forms a complex with UBE3A and Arc during polyubiquitination. In addition, overexpressing Arc enhances the interaction between BECN1 and UBE3A (**Fig. S8D**), further supporting the possibility of an Arc-BECN1-UBE3A complex. Together, these data suggest that BECN1 serves as an adaptor to recruit Arc to UBE3A for ubiquitination.

### BECN1 promotes Arc polyubiquitination and proteasomal degradation via binding the E3 UBE3A

Arc is encoded by an immediate-early gene and regulates glutamatergic synaptic plasticity by mediating endocytosis of AMPARs. In normal excitatory neurons, upon neuronal stimulation, Arc protein is induced and promotes endocytosis of AMPARs, leading to a transient reduction in synaptic activity (*36–38*); several hours post neuronal stimulation, UBE3A polyubiquitinates Arc and promotes proteasomal degradation of Arc (*39*), preventing further endocytosis of AMPARs. Thus, we speculated that Arc is a link by which BECN1 regulates synaptic cell-surface AMPARs. To determine whether BECN1 is important for Arc polyubiquitination, we analyzed the level of ubiquitinated Arc among all polyubiquitinated proteins pulled down by Tandem Ubiquitin Binding Entities (TUBE) beads in the hippocampus of global *Becn1*^+/-^ KO mice and WT littermate mice. We found that *Becn1*^+/-^ KO mice have lower ubiquitinated Arc levels than control mice (**Fig. 5A**), suggesting defective Arc polyubiquitination upon loss of BECN1 in vivo. In line with this finding, Arc protein is stabilized in the *Becn1*^+/-^ KO mouse hippocampus (**Fig. 5B**), while hippocampal mRNA transcription of Arc is unaltered in either *Becn1*^+/-^ KO or CamKII^Becn1^-KO mice (**Fig. S9A**), supporting a role of BECN1 in regulating Arc protein stability. In addition, we found that *BECN1* KO cells show higher levels of Arc protein than WT cells, and a time course of proteasomal inhibitor MG132 treatment restores Arc protein levels in WT cells to a similar level as in *BECN1* KO cells (**Fig. 5C**, **Fig. S9B**), suggesting that KO of *BECN1* stabilizes Arc via blocking the ubiquitin-proteasomal pathway. Thus, we conclude that BECN1 promotes polyubiquitination and proteasomal degradation of Arc.

**Figure 5.**
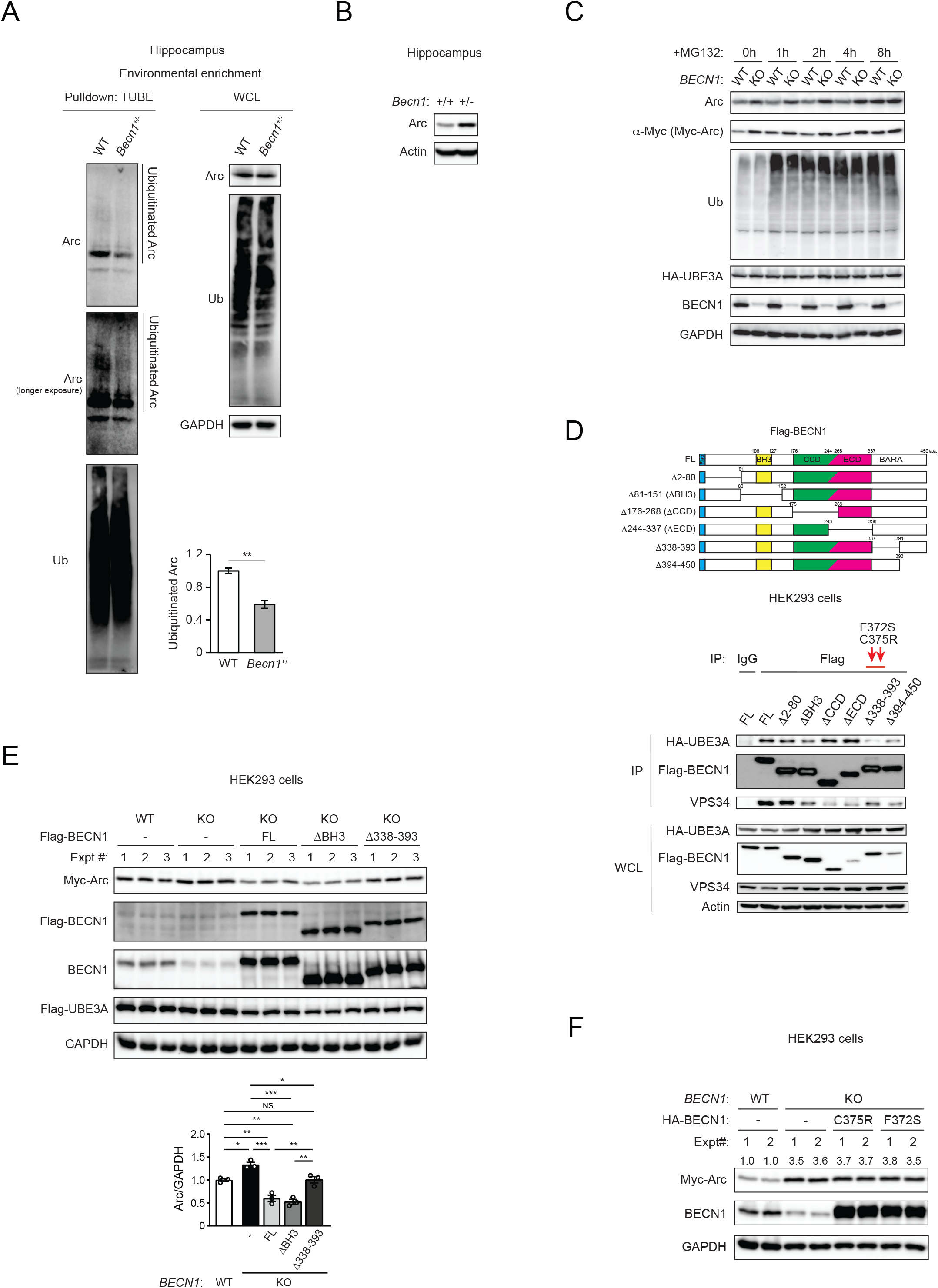
BECN1 promotes Arc polyubiquitination and proteasomal degradation via binding with UBE3A. **(A)** WB and quantification of levels of Arc protein via TUBE pulldown from hippocampal lysates of WT and *Becn1*^+/-^ KO mice subject to 4 h of environmental enrichment. N=3. T-test. **(B)** WB analysis of Arc in the hippocampus of WT (+/+) or *Becn1*^+/-^ KO (+/-) mice. **(C)** WB analysis of Arc levels in WT and *BECN1* CRISPR KO HEK293 cells transfected with Myc-Arc and HA-UBE3A and treated with the proteasomal inhibitor MG132 (10 μM) for the indicated time course (0-8 h). **(D)** C-terminal amino acids 338-393 of BECN1 are required for BECN1-UBE3A interaction. Co-IP of HA-UBE3A by full-length (FL) and indicated deletion mutants of Flag-BECN1 in HEK293 cells. The known BECN1-binding protein VPS34 serves as a positive control. Red arrows indicate the location of BECN1^F372S^ and BECN1^C375R^ variants. BH3: Bcl-2 Homology 3 domain; CCD: Coiled-Coil Domain; ECD: Evolutionarily Conserved Domain; BARA: β-α repeated autophagy-specific domain. **(E)** WB analysis and quantification of Arc protein levels in WT and *BECN1* KO HEK293 cells transfected with Flag-UBE3A, Myc-Arc, and full-length (FL) BECN1, BECN1^ΔBH3^ or BECN1^Δ338-393^, as well as an empty vector as negative control. Cells were treated with 10 μM MG132 for 4 h prior to sample collection. **(F)** WB of Arc protein levels in WT and *BECN1* KO HEK293 cells transfected with Myc-Arc and an empty vector, BECN1^F372S^ or BECN1^C375R^. Cells were treated with 10 μM MG132 for 4 h prior to sample collection. Intensity quantification is listed above each band. *, P<0.05; **, P<0.01; ***, P<0.001; NS, not significant. All error bars represent mean ± SEM.

BECN1 has 3 key domains: the BH3 domain (BCL2 binding domain) within an intrinsically disordered N-terminal region, the coiled-coil domain (CCD) for protein interactions, and the C-terminal BARA domain that contains an evolutionarily conserved domain (ECD) for membrane binding (*2, 40*). To reveal the physiological function of the BECN1-UBE3A interaction, we generated deletion mutants of BECN1 lacking the N-terminal intrinsically disordered region, the BH3 domain, the CCD, the ECD, or the C-terminal BARA regions, respectively (**Fig. 5D**). Via co-IP, we found that the BECN1^Δ338-393^ mutant dramatically decreases BECN1-UBE3A interaction, while partially maintaining its binding with the autophagy kinase VPS34 (**Fig. 5D**), suggesting that amino acids 338-393 in the BARA domain of BECN1 are important for UBE3A binding. Further, full-length (FL) BECN1, or the BECN1^ΔBH3^ mutant that maintains BECN1-UBE3A binding, was able to reduce Arc levels in *BECN1* KO cells, but BECN1^Δ338-393^ failed to do so (**Fig. 5E**), suggesting that BECN1-UBE3A binding is essential for maintaining Arc at a normal low level. Importantly, the two missense BECN1 variants in affected humans, F372S and C375R, both of which are located within the 338-393 amino acids region, failed to rescue Arc protein levels in *BECN1* KO cells (**Fig. 5F**), supporting the conclusion that excess Arc accumulation is a disease-causing mechanism underlying human *BECN1* variant-associated neurodevelopmental diseases. Overall, these data demonstrate that BECN1 regulates Arc ubiquitination and proteasomal degradation by interacting with UBE3A.

### Peptide blockade of Arc-induced AMPAR endocytosis rescues abnormal social interaction in *Becn1* mutant mice

Tat-GluR23Y is a cell-permeable peptide that contains the tyrosine-based trafficking signal (869YKEGYNVYG877) required for Arc-mediated AMPAR endocytosis, and thus functions as a specific competitive blocker of Arc-induced AMPAR endocytosis (*41, 42*). We found that Tat-GluR23Y treatment (3 μmol/kg) rescues both sociality and social novelty of CA3^Becn1^-KO mice in three-chamber tests (**Fig. 6A**), suggesting that social interaction defects caused by loss of BECN1 in CA3 neurons can be reversed by blocking AMPAR endocytosis. Thus, these data support that BECN1 maintains normal social development via preventing Arc-induced AMPAR endocytosis in hippocampal CA3 neurons.

**Figure 6.**
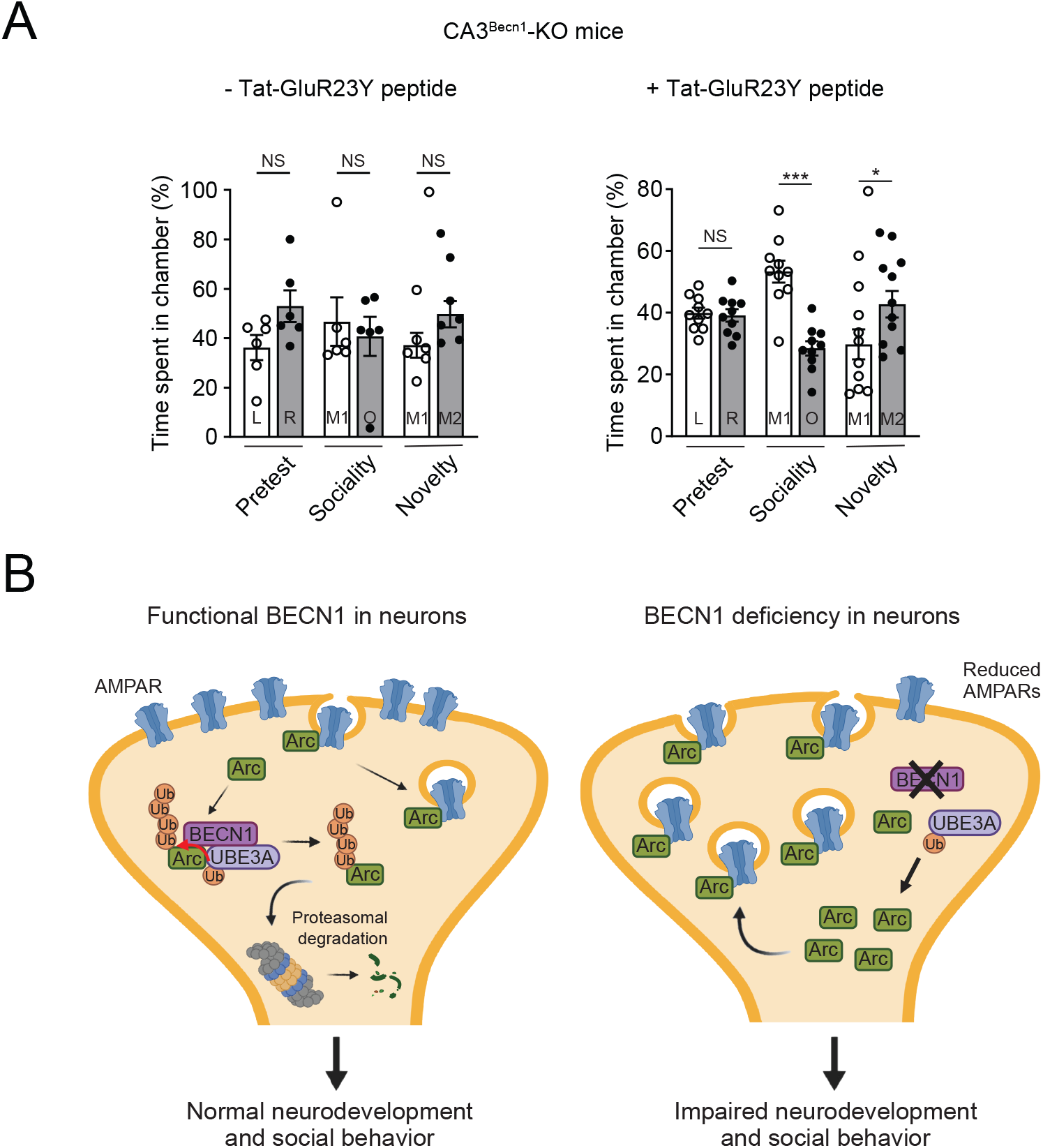
Blocking Arc-mediated AMPAR endocytosis rescues abnormal social behavior induced by loss of BECN1 in CA3 neurons. **(A)** Three-chamber test of CA3^Becn1^-KO mice with and without i.p. injection of Tat-GluR23Y peptides once per day for 4 days. N=6-10. Two-way ANOVA with Bonferroni test. *, P<0.05; ***, P<0.001; NS, not significant. All error bars represent mean ± SEM. **(B)** Schematic model demonstrating that BECN1 is essential for neural and social development, by recruiting Arc to the E3 ligase UBE3A for Arc polyubiquitination and degradation and thus maintaining postsynaptic AMPAR levels and neurotransmission. In BECN1-deficient neurons, UBE3A-mediated Arc polyubquitination is less efficient, leading to accumulation of Arc, excess endocytosis of AMPARs by Arc, abnormal excitatory neurotransmission, and impaired neurodevelopment and social behavior.

Overall, we propose a model of BECN1-regulated neural and social development in the brain (**Fig. 6B**). In healthy neurons, BECN1 serves as an adaptor to recruit the substrate Arc to the E3 UBE3A for Arc polyubiquitination and degradation, thereby maintaining postsynaptic AMPAR stability, AMPAR neurotransmission, and normal neurodevelopment and social interaction. In BECN1-deficient neurons, UBE3A-mediated Arc polyubiquitination is less efficient, leading to accumulation of Arc, reduced AMPAR cell-surface availability and neurotransmission, and neurodevelopmental and social disorders in humans and animal models.

## Discussion

Here we report that BECN1 is a regulator of neurodevelopment, as evidenced by rare deleterious biallelic *BECN1* variants in four affected children and by genetic studies in animal models, demonstrating a new role for this autophagy protein. The affected individuals showed overlapping neurodevelopmental symptoms, including aberrant neuroanatomy, abnormal speech and language delay, and we used social interaction as a behavioral readout in the mouse studies. Although the BECN1 residues altered in the affected individuals are conserved across multiple species, except in *C. elegans* and yeast, the missense variants do not detectably affect autophagy machinery binding and autophagy activity in cell lines. In addition, BECN1 stabilizes, rather than degrades, postsynaptic AMPARs in neurons, further supporting a non-canonical role of BECN1 in neuronal function.

Our data demonstrate that BECN1 maintains AMPAR levels and neurotransmission by binding both the E3 UBE3A and its substrate Arc, promoting Arc ubiquitination and proteasomal degradation, and preventing Arc-dependent AMPAR internalization. AMPAR is a crucial regulator of synaptic plasticity, and its stability at the postsynaptic cell surface is key for the development of intellectual and social abilities (*43*). Synaptic AMPARs (such as the GluA2 subunit) are either reduced or dysfunctional in autistic mouse models (*39, 44–49*), and in the brains of patients with ASD and neurodevelopmental symptoms (*50, 51*). These findings suggest that the machinery that regulates AMPAR cell-surface trafficking is a vulnerable pathway in ASD and neurodevelopmental disorders. Our findings are helpful to understand how the cell-surface trafficking and stability of AMPARs are regulated, and how such regulation determines neural development and social output.

We acknowledge the limitations of comparing neurodevelopmental data in affected humans to the zebrafish and mouse models reported in this study. For example, neuronal defects induced by inherited biallelic *BECN1* variants (including brain structural changes) are generally more severe than phenotypes in *Becn1* mutant mice. Across-model differences are likely due to the presence of one copy of WT *Becn1* in global *Becn1*^+/-^ KO mice, tamoxifen-inducible *Becn1* KO during adulthood (over 8 weeks of age) in CamKII^Becn1^-KO mice (WT *Becn1* is present during early development), and *Becn1* KO only in specific neuronal populations in CA3^Becn1^-KO mice (WT *Becn1* is present in the rest of brain). In the future, it will be important to leverage the *Becn1*^flox/flox^ mice to determine whether and how BECN1 regulates neuronal and social development at different stages in life, for example, youth/early adolescence (4-5-week, Postnatal day (P) P30-35) vs. young adult (P60) vs. mature adult, given 4 weeks of age in mice is equivalent to early adolescence (human year 10-20 = rodent day 28-56). Notably, *Becn1*^+/-^ KO mice do not have structural defects in the brain, but showed abnormalities in electrophysiology and social interaction. Yet carrier parents of both families are reportedly healthy with no history of neurodevelopmental disease. This difference could be due to multiple factors including species-specific differences and complexity in neural circuitry, or expression levels of the WT vs. variant allele in carrier parents such that a biological or behavioral threshold of disease is not reached as in animal assays. In addition, as observed in many rare Mendelian diseases, there are also phenotypic differences among the four patients in the two families (death at infancy vs. neurodevelopmental defects into adulthood). Though it is challenging to fully understand this question across only four patients in two families, we reasoned that it could be due to additional sensitizers in the genomes (genetic background), or different strengths of causal variants in *BECN1* (nonsense plus missense for the postnatal lethal babies compared to two missense variants in the patients that survived with likely more residual function of BECN1 protein).

This study also reveals the spatial function of BECN1 in neurons. Our findings suggest the involvement of a hippocampal CA3 neural circuitry in the maintenance of social behavior and AMPAR neurotransmission, which is a poorly understood neuronal population in neurodevelopment. Future work includes mapping the upstream brain regions and neuronal populations that process and pass down developmental information to the CA3 neurons. Finally, of note, the cause of death of affected family 1 siblings (compound heterozygotes with one BECN1 nonsense variant and one BECN1^C375R^ missense variant) is poor respiratory effort, raising the possibility that in addition to the hippocampus, BECN1 may regulate AMPAR excitatory neurotransmission in broader brain regions (such as the respiratory center, ventricles and corpus callosum), which is important for overall synapse and brain development and deserves further investigation.

## Supplementary figure legends

**Figure S1.**
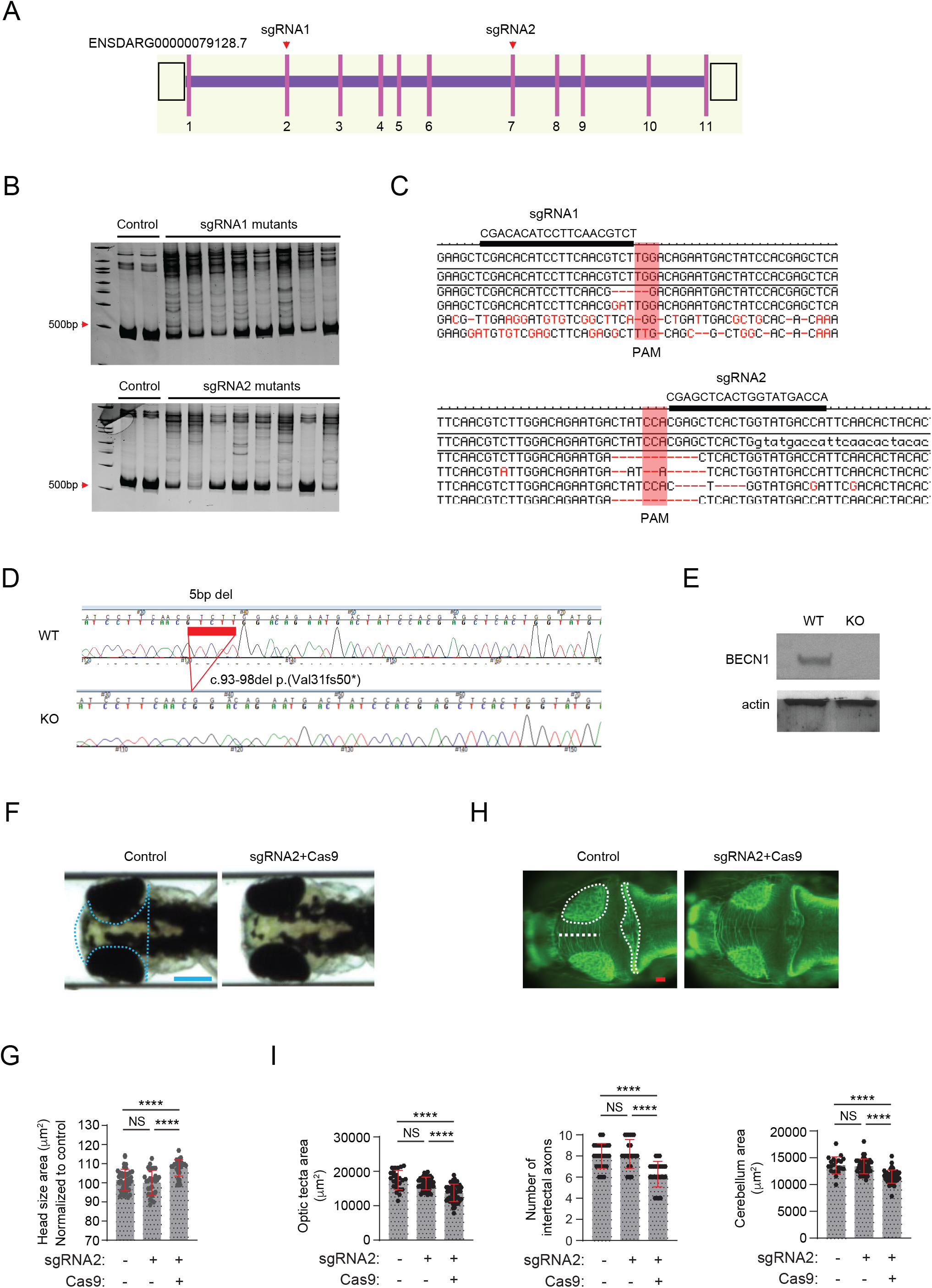
Efficiency and effects of CRISPR/Cas9-mediated *becn1* targeting on neuroanatomy in zebrafish larvae. (**A**) Schematic of the *becn1* locus (GRCz11, *D. rerio*). Pink lines, coding exons; unfilled rectangles, untranslated regions; purple lines, introns. Target position of sgRNAs used are indicated with arrowheads. (**B**) Polyacrylamide gel images showing heteroduplex analysis of two control embryos and eight F0 embryos injected with sgRNA plus Cas9 protein (one lane per embryo). (**C**) Sequence confirmation of insertion/deletions at the target region induced with sgRNAs; PAM, protospacer adjacent motif (red transparent box); mosaicism estimated for each guide is 78% (sgRNA1) and 80% (sgRNA2). (**D**) Sequence chromatograms show representative genotyping results (flanking PCR and sequencing) of WT and *becn1* homozygous mutants. (**E**) Representative immunoblots to detect Becn1 levels in protein lysates from WT and *becn1* homozygous mutant larvae. Actin was used as a loading control. (**F**) Representative bright field dorsal head images acquired at 3 dpf for F0 mosaic animals injected with sgRNA2+Cas9. Scale bar, 300 μm. **(G)** Quantification of head size area (see dotted blue line in panel F) in controls and F0 animals at 3 dpf. n=30-35/condition. Similar results were produced in three biological replicates. (**H**) Representative dorsal fluorescent images of 3 dpf larvae stained with acetylated tubulin antibody to highlight neuroanatomy. Scale bar, 50 μm. **(I)** Quantification of the size of optic tecta (white circle), cerebellum (white elongated area), and number of intertectal axons (crossing the white dotted line). n=40-67/condition. All error bars represent mean ± SEM. ****, P<0.0001; NS, not significant.

**Figure S2.**
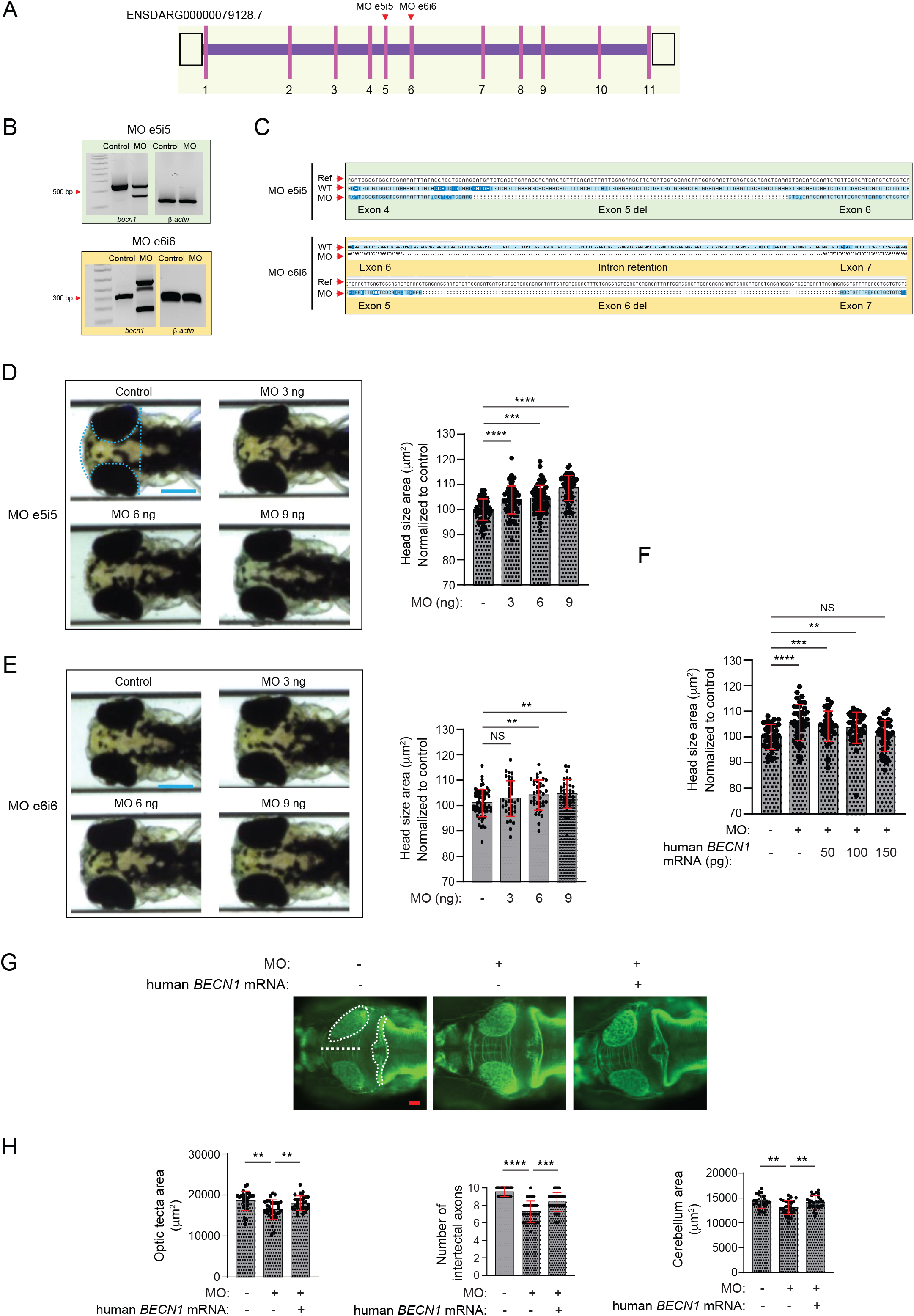
Efficiency and effects of MO-mediated transient *becn1* ablation on neuroanatomy in zebrafish larvae. (**A**) Schematic of the zebrafish *becn1* locus (GRCz11). Pink lines, coding exons; unfilled rectangles, untranslated regions; purple lines, introns. Target position of morpholinos (MOs) used are indicated with arrowheads. (**B**) RT-PCR validation of knockdown efficiency by both MOs. Top panel, agarose gel images showing RT-PCR of *becn1* and *actb* (Δ-actin) from a pool of control embryos or embryos injected with e5i5 MO. Bottom panel, agarose gel images showing RT-PCR of *becn1* and *actb* (Δ-actin) from a pool of control embryos or embryos injected with e6i6 MO. (**C**) Sequence analysis confirms deletion of exon 5 in e5i5 morphants and exon/intron retention in e6i6 morphants. Ref, reference sequence. **(D**) Representative dorsal bright field images (left) and quantification (right) of dorsal head size area (as measured by blue dotted area) in controls or larvae injected with *becn1* (e5i5) MO at different doses (3 ng, 6 ng, and 9 ng). **(E)** Representative dorsal bright field images (left) and quantification (right) of dorsal head size area (as indicated by blue dotted area in panel D) in controls or larvae injected with *becn1* (e6i6) MO at different doses (3 ng, 6 ng, and 9 ng). Scale bar, 300 μm. n=40-97/condition. (**F**) Quantification of dorsal head size area obtained from dorsal bright field images in controls or larvae injected with *becn1* (e5i5) MO with or without increasing doses of WT human *BECN1* mRNA. **(G)** *becn1* morphants show neuroanatomical defects that can be rescued by complementation with WT human *BECN1* mRNA. Representative fluorescent images of 3 dpf larvae stained with anti-acetylated tubulin antibody to highlight zebrafish brain structure in controls or larvae injected with *becn1* (e5i5) MO with or without 150 pg human WT *BECN1* mRNA. Intertectal axons are counted as they cross the midline (demarcated with white vertical line); area of the optic tecta (dashed white circle); and cerebellum (white dashed elongated area). Scale bar, 50 μm. (**H**) Quantification of optic tecta size, intertectal neuron number, and size of cerebellum in panel G. n=25-37/condition. **, P<0.01; ***, P<0.001; ****, P<0.0001; NS, not significant. All error bars represent mean ± SEM.

**Figure S3.**
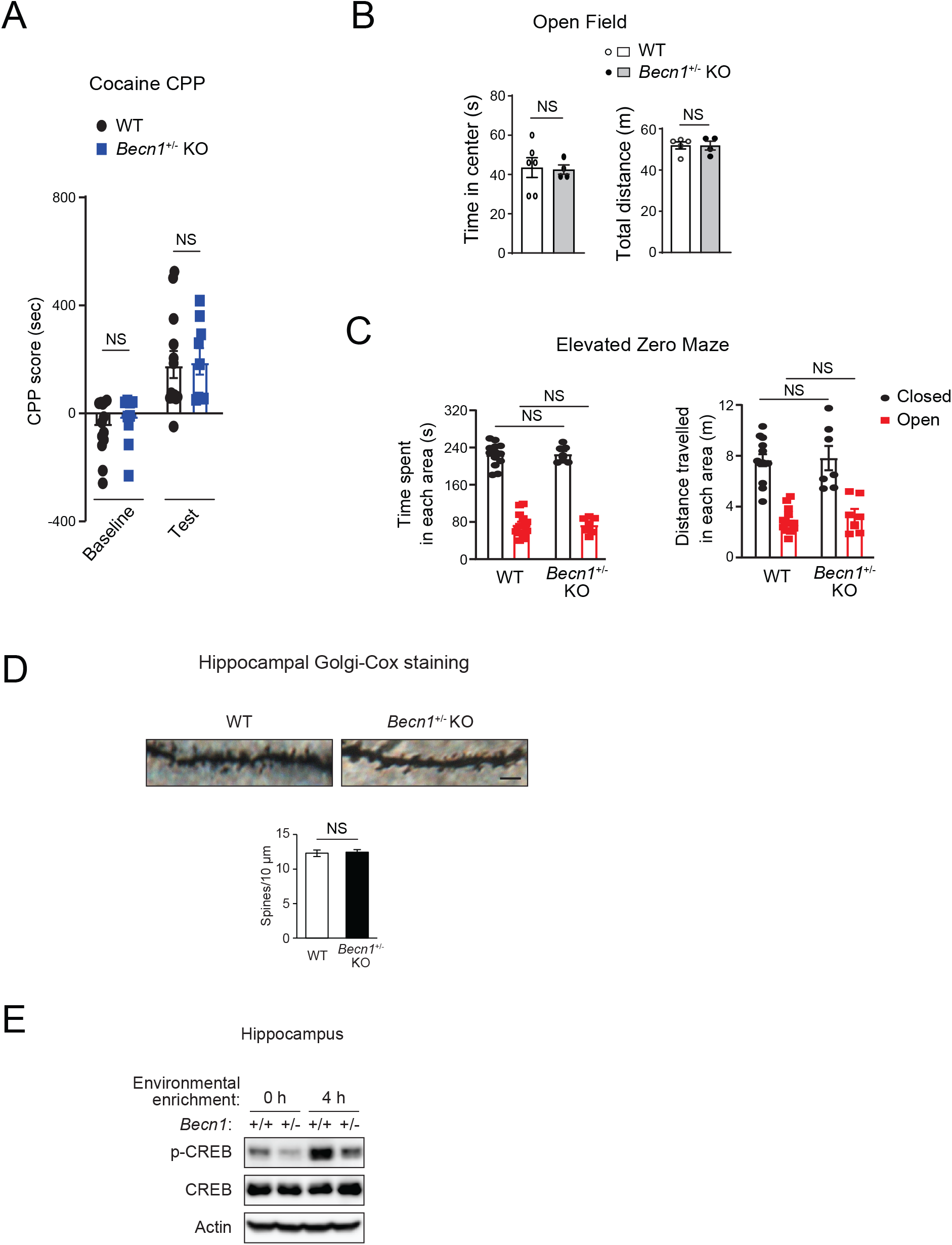
Global haplodeletion of *Becn1* in mice affects neuronal stimulation (p-CREB), but not drug reward, anxiety, or spine pruning. **(A)** Comparable cocaine conditioned place preference (CPP) in WT and *Becn1*^+/-^ KO mice. Mice received alternate cocaine and PBS injection for 3 days (conditioning), and their CPP score (the difference in time spent in the paired cocaine-injecting chamber and the unpaired PBS-injecting chamber) was recorded before (Baseline) and after conditioning (Test). WT, N=14; *Becn1*^+/-^ KO, N=11. **(B)** Comparable time spent by WT and *Becn1*^+/-^ KO mice in the center region (left) and total distance travelled (right) in the open field. N=4-6. **(C)** Comparable time spent (left) and distance travelled (right) by WT and *Becn1*^+/-^ KO mice in the closed or open compartment of the elevated zero maze. WT, N=13; *Becn1*^+/-^ KO, N=7. **(D)** Morphology and quantification of hippocampal neuronal spines in WT and *Becn1*^+/-^ KO mice by Golgi-Cox staining. Scale bar, 2.5 µm. N=21 areas from 3 mice. **(E)** WB analysis of phosphorylated CREB (p-CREB) in the hippocampus of WT or *Becn1*^+/-^ KO mice with or without 4 h of environmental enrichment. T-test. NS, not significant. All values represent mean ± SEM.

**Figure S4.**
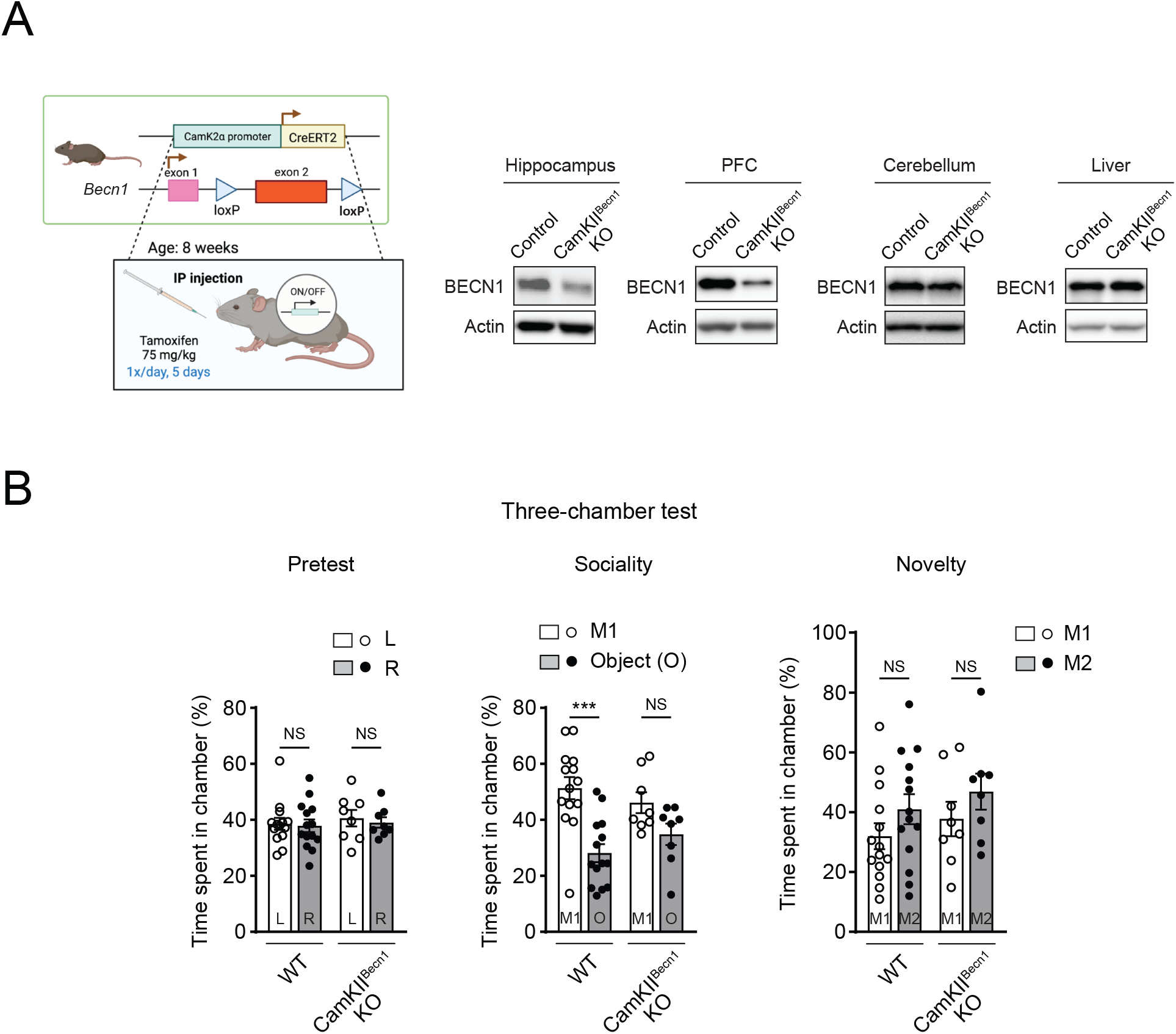
Generation and characterization of inducible CamKII^Becn1^-KO mice. **(A)** Generation of inducible CamKII^Becn1^-KO mice. Depletion of BECN1 in the hippocampus and prefrontal cortex (PFC), but not the cerebellum or liver, of CamKII^Becn1^-KO mice (tamoxifen-injected *Becn1*^flox/flox^; CamKIIɑ-Cre/ERT2 mice) vs. control mice (tamoxifen-injected *Becn1*^floxlflox^ mice), analyzed by WB. Cre expression is induced by i.p. injection of tamoxifen (75 mg/kg body weight) once a day for 5 days at 8 weeks of age. **(B)** *Becn1* KO in CamKII neurons during adulthood impairs social interaction. Three-chamber test of male CamKII^Becn1^-KO mice and control mice 3 weeks after tamoxifen injection. N=8-14. Two-way ANOVA with Bonferroni test. ***, P < 0.001; NS, not significant. All values represent mean ± SEM.

**Figure S5.**
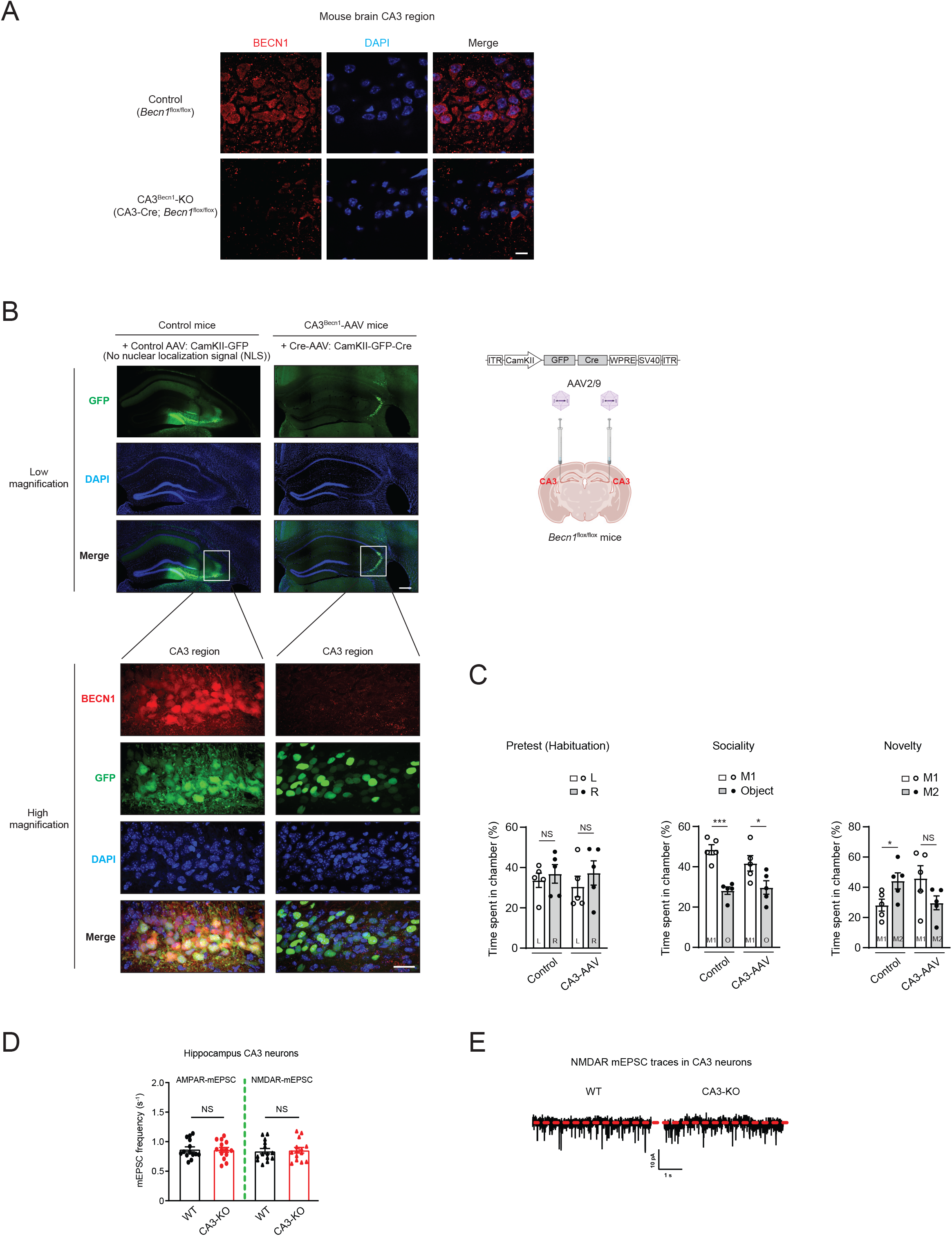
Generation and characterization of two CA3 neuron-specific *Becn1* KO mouse models (genetic and AAV-mediated KO mice). **(A)** Validation of BECN1 depletion in the genetic model CA3^Becn1^-KO mice, resuling from *Becn1*^flox/flox^ mice crossed with CA3-Cre (Grik4-Cre/G32-4 Cre) mice. Immunofluoresence microscopy of BECN1 in the hippocampal CA3 region of control (*Becn1*^flox/flox^) and CA3^Becn1^-KO mice. Scale bar, 10 µm. **(B)** Generation and validation of AAV-mediated CA3 neuron-specific *Becn1* KO mice (CA3^Becn1^-AAV mice). Immunofluoresence microscopy of BECN1 in CA3 neurons of *Becn1*^flox/flox^ mice that received bilateral stereotaxic microinjection of control AAV (AAV2/9-CamKII-GFP) or Cre-AAV (AAV2/9-CamKII-GFP-Cre) into the CA3 region of both hemispheres. Low magnification: scale bar, 250 µm; high magnification: scale bar, 25 µm. **(C)** Three-chamber test of control and CA3^Becn1^-AAV mice demonstrating reduced sociality and social novelty in CA3^Becn1^-AAV mice. N=5. T-test. **(D)** Frequency of AMPAR-dependent mEPSC and NMDAR-dependent mEPSC in hippocampal CA3 neurons of WT and CA3^Becn1^-KO mice. **(E)** Representative traces of NMDAR mEPSC in hippocampal CA3 neurons of WT and CA3^Becn1^-KO mice.

**Figure S6.**
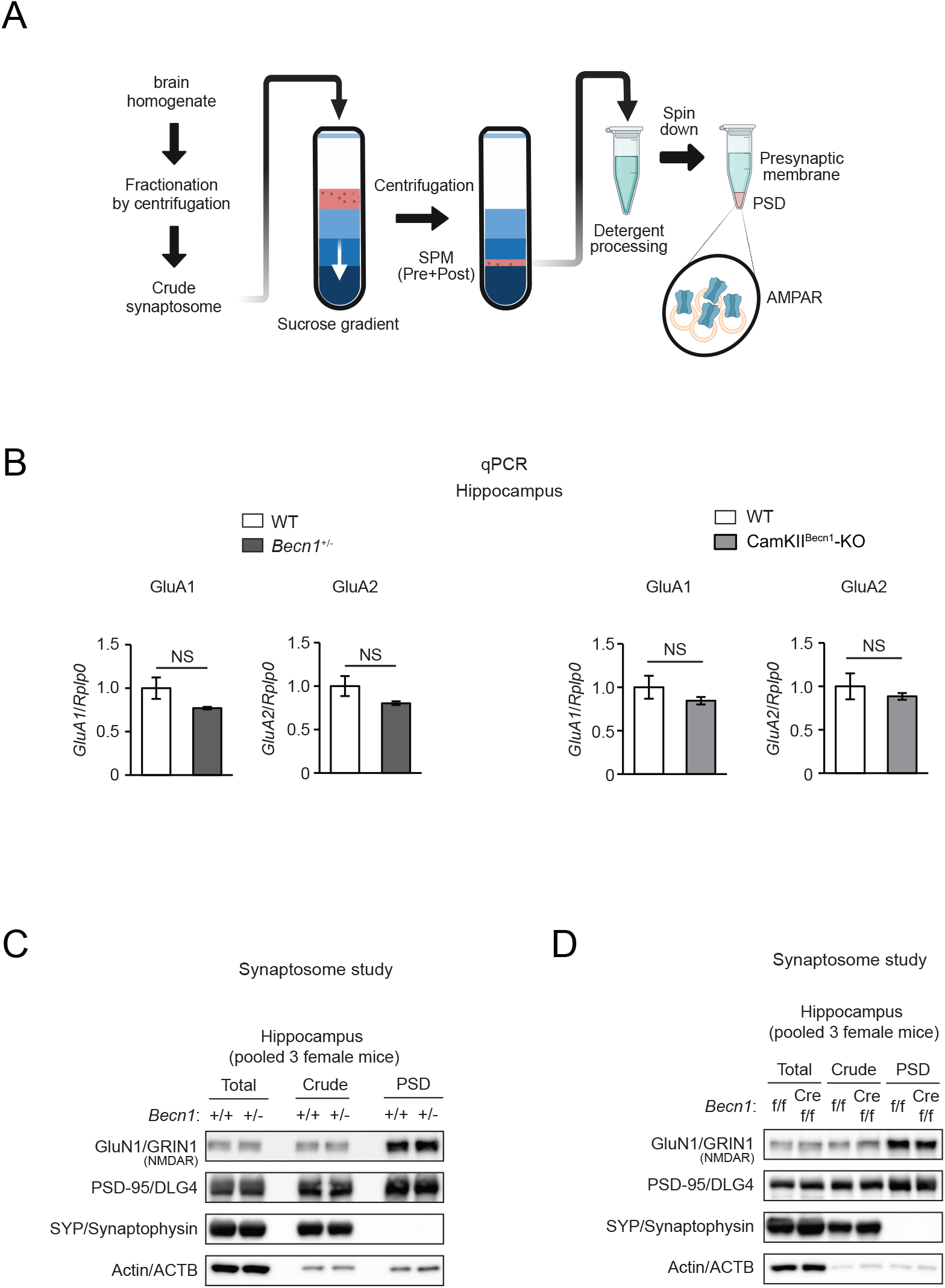
*Becn1* KO does not affect mRNA levels of AMPAR subunits, or postsynaptic protein levels of NMDAR. **(A)** Schematic method of isolating presynaptic and postsynaptic membrane fractions from mouse brain. **(B)** qPCR analysis of AMPAR subunits GluA1 and GluA2 mRNA expression in the hippocampus of control, *Becn1*^+/-^ KO mice, and CamKII^Becn1^-KO mice. N=5-6. T-test. NS, not significant. All values represent mean ± SEM. **(C)** Synaptosome studies analyzed by WB on the NMDAR subunit GluN1 in the hippocampus pooled from 3 *Becn1*^+/-^ KO (+/-) mice and 3 WT littermates (+/+). **(D)** Synaptosome studies analyzed by WB on the NMDAR subunit GluN1 in the hippocampus pooled from 3 CamKII^Becn1^-KO (Cre f/f) mice and 3 control (f/f) mice (tamoxifen-injected *Becn1*^floxlflox^ mice). Total, total lysates; Crude, crude synaptosomes; PSD, postsynaptic density; SYP/Synaptophysin, presynaptic marker; PSD95, postsynaptic marker.

**Figure S7.**
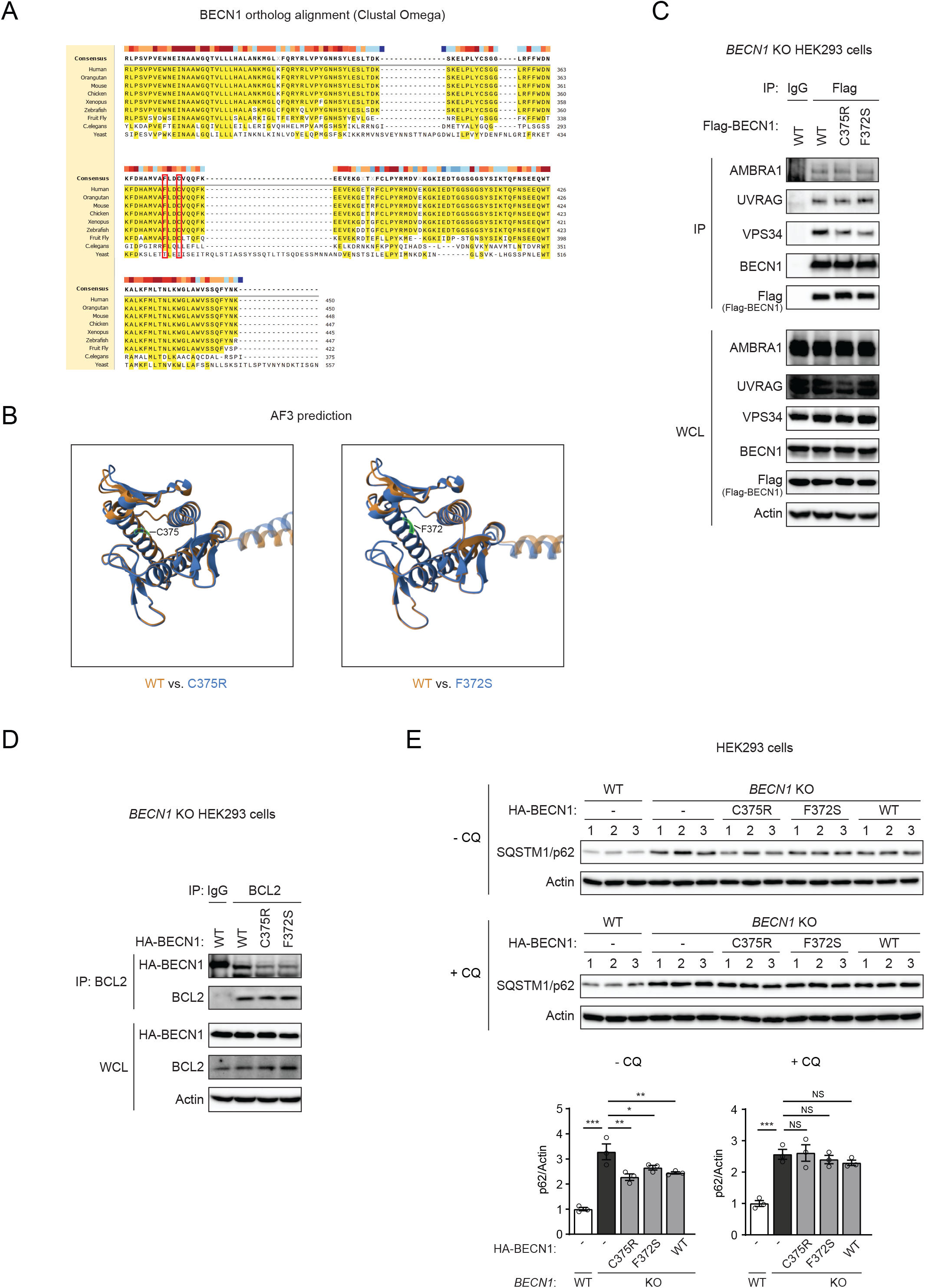
BECN1 F372S and C375R missense variants maintain BECN1 protein structure, autophagy and endocytic machinery binding, and autophagy activity of BECN1. **(A)** Amino acid alignment of BECN1 C-terminus across species. Positions of variants identified in affected humans are marked with red boxes. **(B)** Structure of C-terminal WT BECN1 and BECN1 variants identified in this study predicted by AlphaFold3 (AF3). **(C)** Co-IP of known BECN1-binding partners in the autophagy and endocytic machineries (AMBRA1, UVRAG, and VPS34) with WT BECN1, or variants BECN1^C375R^ and BECN1^F372S^ in *BECN1* KO HEK293 cells transfected with Flag-BECN1 plasmids. IgG IP serves as a negative control. **(D)** Co-IP of WT BECN1, or variants BECN1^C375R^ and BECN1^F372S^, with endogenous BCL2 in *BECN1* KO HEK293 cells transfected with HA-BECN1 plasmids. IgG IP serves as a negative control. **(E)** WB and quantification of the autophagy cargo receptor SQSTM1/p62 in WT cells, or *BECN1* KO HEK293 cells transfected with vector, BECN1^C375R^, BECN1^F372S^ or WT BECN1. Cells were treated with or without 10 μM lysosomal inhibitor chloroquine (CQ) for 4 h prior to sample collection. N=3. One-way ANOVA with Dunnett’s test. *, P<0.05; **, P<0.01; ***, P<0.001; NS, not significant. All values represent mean ± SEM.

**Figure S8.**
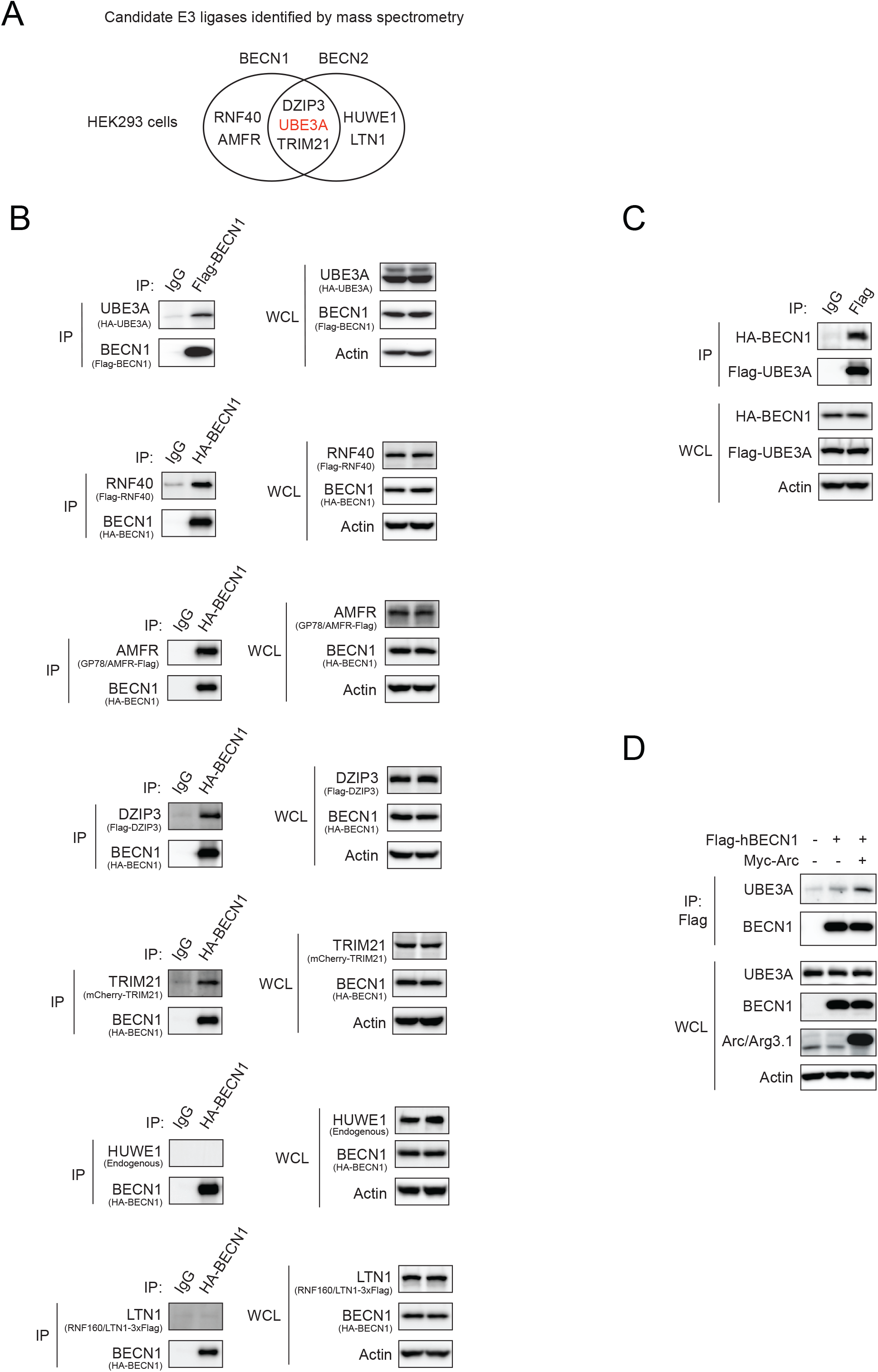
Interaction between BECN1 and the E3 ligase UBE3A. **(A)** Top E3 ligase hits by IP-MS analysis for BECN1- or BECN2-interacting proteins in HEK293 cells stably expressing Flag-BECN1 or Flag-BECN2. **(B)** Co-IP validation of candidate BECN1-binding E3 ligases in HEK293 cells identified by IP-MS in (A). Co-IP of UBE3A by BECN1 validated the interaction between BECN1 and UBE3A. **(C)** Co-IP of BECN1 by UBE3A in HEK293 cells transfected with HA-BECN1 and Flag-UBE3A. **(D)** Co-IP of endogenous UBE3A by Flag-BECN1 in the presence or absence of Myc-Arc expression in HEK293 cells.

**Figure S9.**
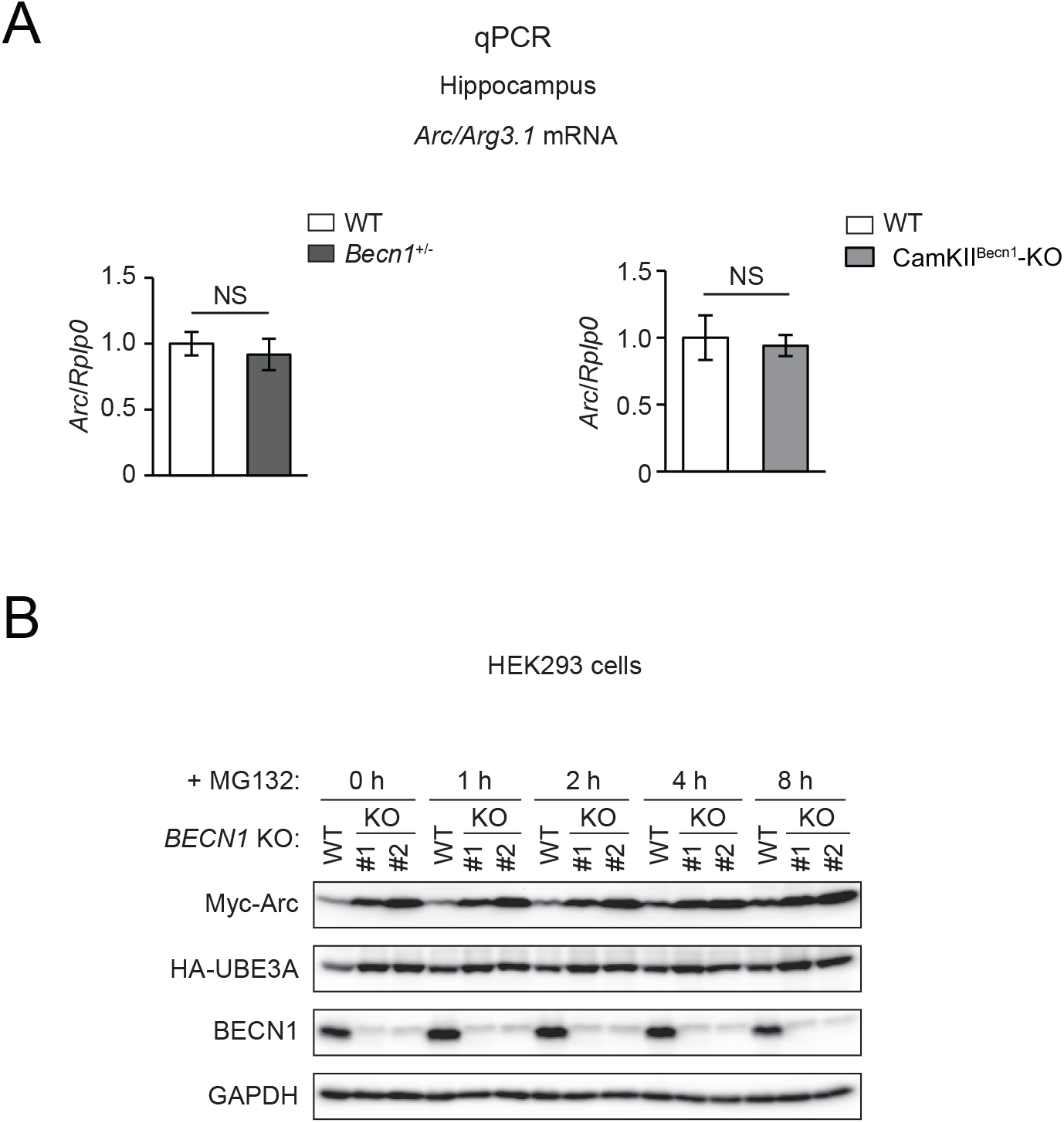
*BECN1* KO stabilizes Arc via blocking the proteasomal pathway. **(A)** qPCR analysis of *Arc* mRNA in the hippocampus of control, *Becn1*^+/-^ KO mice, and CamKII^Becn1^-KO mice. N=5-6. T-test. NS, not significant. All values represent mean ± SEM. **(B)** WB analysis of Arc protein levels in WT HEK293 cells and two *BECN1* CRISPR KO clones expressing Myc-Arc and HA-UBE3A treated with MG132 (10 μM) for the indicated time course (0-8 h).

## Supplementary Information

### Supplementary case reports

#### Family 1 (AC0001)

The Family 1 parents are nonconsanguineous; both report an unremarkable family history and have a healthy son. The family was enrolled in genetics research, and we generated ES data from both affected siblings and their parents; bioinformatic filtering and Sanger confirmation of candidates confirmed the *BECN1* alleles while not identifying any other plausible candidate causal variants. Based on the medRxiv policy on identifying information, we advise that readers contact the corresponding author for access to medical history data details.

#### Family 2 (19DG)

Individual 2-IV-1 (19DG0859) is an affected female who had severe intellectual disability with abnormal behavior and communication, and neurodevelopmental delay. Her metabolic workup was unremarkable. The younger sister, 2-IV-2 (19DG0937) was diagnosed with neurodevelopmental delay and intellectual disability. Based on the medRxiv policy on identifying information, we advise that readers contact the corresponding author for access to medical history data details.

## Materials and methods

### Ascertainment of affected individuals

Clinical assessment of affected individuals was performed by local specialists at each participating medical center. Affected individuals were referred for research because they met inclusion criteria of an undiagnosed likely genetic condition that involved impaired neurodevelopment. Each research study was approved by local ethics committees, including the Institutional Review Board at Lurie Children’s Hospital (IRB2019-3084) and King Faisal Specialist Hospital and Research Center (RAC# 2121053). Written informed consent for participation in research was obtained from healthy parents and (or on behalf of) their affected children. Clinical imaging (prenatal ultrasound, magnetic resonance imaging [MRI]) was performed as part of standard clinical care. GeneMatcher (*54*) was used to assemble a cohort of families harboring biallelic *BECN1* variants.

### Genetic analyses

#### Family 1

Clinical genetic testing was performed on Family 1 as standard of care and included mother-proband duo ES (GeneDx) and genome-wide SNP copy number microarray (Lurie Children’s Hospital; Infinium Global Diversity Array, Illumina). Residual genomic DNA was obtained for research-based parent-proband quad ES (LC Sciences, LLC) and analysis (Varsome Clinical version 11.8) as described (*55*). Briefly, the Agilent SureSelect Human All Exon V6 kit was used for exome capture and paired-end 150 bp sequencing reads were generated on an Illumina Novaseq6000. After low-quality reads were removed, the remaining reads were mapped to the human genome reference (UCSC GRCh37/hg19). Variants were called using the Genome Analysis Tool Kit (GATK, version 3.3). Post QC data were filtered to retain functional variants shared exclusively between the affected individuals, including changes impacting amino acid sequence or mRNA splicing. Resulting variants with minor allele frequency (MAF) ≤ 1% in public SNP databases (Genome aggregation database [gnomAD]) were retained for further evaluation. Sanger sequencing was performed for secondary confirmation of *BECN1* variants and segregation analysis.

#### Family 2

Genome-wide genotyping was performed on all available family members using the Axiom platform following the manufacturer’s protocol. Regions of homozygosity (ROH) measuring at least 2 Mb were identified and used as indicators of autozygosity through the autoSNPa tool (https://dna-leeds.co.uk/autosnpa/). ES, along with filtering of candidate variants based on the defined autozygome intervals, was performed as described (*56*).

### Zebrafish ethics approval and husbandry

All zebrafish experiments were performed according to protocols approved by the Northwestern University Institutional Care and Use Committee (IACUC). Embryos were obtained by natural mating of adult WT zebrafish (NIH line). Embryos were grown in egg water (0.3 g/L NaCl, 75 mg/L CaSO4, 37.5 mg/L NaHC03, 0.003% methylene blue) at 28°C until specific phenotypic endpoints.

### Mice

All mouse care and experimental procedures were conducted in accordance with protocols approved by the Northwestern University Institutional Animal Care and Use Committee (IACUC). Mice were housed under a 14 h/10 h light/dark cycle with ad libitum access to chow diet and water. C57BL/6J (JAX #000664), *Becn1*^flox^ (JAX #028794), Camk2a-Cre/ERT2 (JAX #012362), and CA3-Cre (Grik4-Cre/G32-4 Cre mice, JAX #006474) mice were obtained from The Jackson Laboratory. The *Becn1*^+/−^ knockout mouse line was described previously (*6*). All experiments were performed using sex- and age-matched mice.

To induce Cre-mediated recombination, 6–8-week-old mice were intraperitoneally injected with tamoxifen (APExBIO, #B5965), dissolved in corn oil (MilliporeSigma, #C8267), at a dose of 75 mg/kg for 5 consecutive days.

Environmental enrichment cages were assembled as shown in Fig. 2G, following a previously described protocol (*57*). Mice were housed in the environmental enrichment cages for 4 hours.

### CRISPR-Cas9 genome editing in zebrafish

Reciprocal BLAST searches of human BECN1 (NP_001300927.1) were used against the translated zebrafish genome to confirm a single zebrafish ortholog (GRCz11 Ensembl ID: ENSDART00000115237.5). Two single guide (sg)RNAs targeting *becn1* were designed using CHOPCHOPv2 (*58*). With a synthetic oligo template, sgRNAs were generated with the GeneArt precision gRNA kit (Thermo Fisher Scientific). One nl of cocktail containing either sgRNA alone (100 pg) or sgRNA plus Cas9 (200 pg, PNA Bio) was microinjected into one-cell-staged zebrafish embryos and allowed them to grow to 2 days post-fertilization (dpf). To characterize mosaicism in F0 mutants, genomic DNA was extracted from individual embryos (n=2 controls and n=8 sgRNA+Cas9-injected) and the target region of each sgRNA was PCR-amplified with flanking primers. Heteroduplex analysis was performed on PCR products; they were denatured and reannealed slowly, loaded on a 16% polyacrylamide gel (Thermo Fisher Scientific) and allowed to migrate initially for 5 minutes at 120 V and then for 2 hours at 150 V prior to imaging on a ChemiDoc (BioRad). PCR products from 5 embryos were randomly selected, purified and cloned in TOPO-TA cloning vector (Thermo Fisher Scientific). Sixteen colonies were miniprepped from each embryo and Sanger-sequenced using BigDye Terminator v3.1 cycle sequencing (Applied Biosystems).

### Characterization of *becn1* germline mutant zebrafish

CRISPR/Cas9 genome-edited F0 *becn1* embryos were reared to adulthood (∼3 months). F0 mutants were outcrossed with WT AB adults and resulting F1 progeny were grown for another 3 months. F1 heterozygous adult animals were genotyped using DNA extracted from fin clip biopsies; the sgRNA target site was PCR-amplified and Sanger sequencing was performed. F1 heterozygous animals harboring a 5 bp deletion (GenBank ID NM_200872.1: c.93_98del; p.(Val31delfs*50) were in-crossed to generate F2 homozygous animals. All experiments with germline mutants were performed in F2 generation sibling animals.

### Transient suppression of *becn1*

Splice blocking morpholinos (MOs) were designed to target *becn1* exon 5 (e5i5) and 6 (e6i6) splice donor sites, respectively (Gene Tools). *becn1* was transiently suppressed by microinjecting 1 nl of MO at different doses (3 ng, 6 ng and 9 ng) into zebrafish embryos staged from one-to-four cells. MO efficiency was validated by extracting total RNA from pools of embryos at 2 dpf (30 animals/condition; controls and MO-injected) using Trizol (Thermo Fisher Scientific) according to manufacturer’s instructions. cDNA was synthesized with the QuantiTect Reverse Transcription kit (QIAGEN). RT-PCR of the MO target site was performed using flanking primers and the amplified PCR product was separated on a 1% agarose gel. The resulting PCR bands were excised with a sterile blade and gel-purified with the QIAquick gel extraction kit (QIAGEN). PCR product was cloned into TOPO-TA cloning vector (Thermo Fisher Scientific) and the plasmids were purified from resulting colonies (n=4/PCR product) and sequenced using BigDye Terminator v3.1 chemistry (Applied Biosystems).

### Zebrafish whole mount immunostaining

Whole mount immunostaining was performed as described (*59*). Dechorionated embryos were fixed overnight in Dent’s solution at 3 dpf (80% MeOH + 20% DMSO for acetylated tubulin). The embryos were rehydrated sequentially with decreasing concentrations of MeOH (75% MeOH/25%PBS; 50% MeOH/50%PBS; 50% MeOH/75%PBS). The embryos were gently washed with bleaching solution (9 mL PBST + 1 mL H2O2 + 0.05 g KOH) for 12-15 minutes and washed 3X with PBST, treated with proteinase K, post fixed with 4% PFA and incubated in blocking buffer for 1 hour. Samples were incubated overnight at 4°C in primary antibody diluted in blocking solution (Acetylated Tubulin antibody, T7451, Sigma-Aldrich, 1:500). The embryos were washed twice and treated with secondary antibody (Alexa Fluor 488 goat anti-rabbit IgG, A-11008, Thermo Fisher Scientific, 1:500) for 2 hours. Subsequently, the embryos were washed twice with immunofluorescence (IF) buffer (1% BSA in PBST) and processed them for imaging. Fluorescent signal was captured with a Nikon AZ100 microscope equipped with a Nikon camera and controlled by Nikon NIS Elements Software (Version 5.20.01).

### Live automated imaging of zebrafish larvae

Images of tricaine-anesthetized larvae at 3 dpf were captured using the Vertebrate Automated Screening Technology (VAST) Bioimager (Union Biometrica) mounted to an AXIO Imager.M2m microscope (Zeiss) with a 10x objective lens. Larvae were passed sequentially through a 600 μm capillary on the detection platform. Each larva was detected and oriented automatically with predetermined templates in the VAST software (version 1.2.6.7); the instrument operated in automatic imaging mode with a 70% minimum similarity threshold, as described (*60*). Dorsal bright field images were captured with the VAST onboard camera.

### Zebrafish image analysis

ImageJ (NIH) was used for quantitative analysis of morphological features of zebrafish embryos and larvae. To assess the head size in larvae at 3 dpf, the area between the eyes and extending to the most anterior point was measured on dorsal bright field images. To evaluate the neuronal organization on acetylated tubulin antibody-stained larvae at 3 dpf, the area of the optic tecta or cerebellum was measured, and the number of axons connecting the two optic tecta at the midline was counted on dorsal images.

### Stereotaxic microinjection of adeno-associated virus (AAV)

Intracranial microinjection of AAV using a stereotaxic apparatus was performed as described (*61*). Seven-week-old mice were bilaterally injected into the hippocampal CA3 region with 150 nL of either AAV2/9-CamKII.HI.GFP-Cre.WPRE.SV40 (5.0 × 10¹² VG/mL; Addgene #105551-AAV9) or AAV2/9-CaMKIIa-EGFP (5.6 × 10¹² VG/mL; Addgene #50469-AAV9). Stereotaxic coordinates relative to bregma were as follows: anteroposterior (AP), −2.06 mm; mediolateral (ML), ±2.35 mm; dorsoventral (DV), −2.35 mm. Four weeks after AAV injection, the mice were used for behavior tests.

### Mouse behavioral tests

The three-chamber social preference test was performed as described (*62*). The three-chamber arena measured approximately 30″ × 12″ × 12″ (length × width × height). The left and right chambers each measured approximately 11″ × 12″, and the center chamber measured 8″ × 12″; chambers were separated by clear Plexiglas partitions. Chambers were cleaned with C-Dox followed by 70% ethanol before each test to eliminate residual odors. Each arena was housed in an individual sound- and light-attenuated chamber equipped with a recording camera and fluorescent lighting connected to an external computer. Mouse behavior was recorded and analyzed using LimeLight software (version 6; Actimetrics).

Test mice were transferred to the behavioral testing room and allowed to habituate for at least 1 hour. Mice were then placed in the center chamber and allowed to habituate for 10 min. The test consisted of three sequential 10-min phases: habituation (pre-test), sociality, and novelty. During the habituation phase, empty inverted pencil cups containing paper balls were placed in both side chambers, and mouse behavior was recorded for 10 min. During the sociality phase, an age- and sex-matched unfamiliar wild-type mouse (M1) was placed under an inverted pencil cup in one side chamber, while a block was placed under an inverted pencil cup in the opposite chamber as an object control (O). During the novelty phase, a novel unfamiliar mouse (M2) was placed under the inverted pencil cup that previously contained the object.

To block AMPA receptor endocytosis in vivo, mice were intraperitoneally injected with TAT-GluR23Y (NovoPro, #318742) at a dose of 3 μmol/kg for 4 consecutive days. The final intraperitoneal injection on day 4 was administered 1 hour before the three-chamber social preference test.

The cocaine conditioned place preference (CPP) test, open field test, and elevated zero maze test were performed as described (*61*). Briefly, for CPP, a randomized unbiased procedure was used. The apparatus consisted of two rectangular compartments (45 cm × 20 cm × 20 cm) separated by a removable door, with distinct visual and tactile cues. One compartment had black-and-white striped walls, while the other had black dots on a white background. Floors were interchangeable and had either apertured or smooth textures. The procedure consisted of three phases: baseline pretesting, conditioning, and testing. During pretesting (day 1), mice were allowed free access to both compartments for 20 min, and time spent in each compartment was recorded using LimeLight software. For cocaine CPP, mice received alternating intraperitoneal injections of cocaine (15 mg/kg) or PBS and were confined to the corresponding chamber for 30 min following each injection. Conditioning consisted of two sessions per day for 4 consecutive days, separated by at least 3 hours. CPP testing was conducted using the same procedure as the pretest. CPP scores were calculated by subtracting the time spent in the PBS chamber from the time spent in the cocaine chamber.

For the open field test, mice were placed in a wooden arena (56 cm × 56 cm), and locomotor activity was recorded for 10 min using LimeLight software. Total distance traveled and time spent in the center zone (20 cm × 20 cm) were quantified. The arena was cleaned with 70% ethanol between trials.

Anxiety-like behavior was assessed using the elevated zero maze. The apparatus consisted of a circular track (5 cm wide, 45 cm inner diameter) elevated 35 cm above the floor, with two open quadrants and two closed quadrants enclosed by 15-cm-high walls. Mice were placed in a closed quadrant and allowed to explore the maze for 5 min. Total distance traveled, distance traveled in open quadrants, and time spent in open quadrants were analyzed using LimeLight software.

### Postsynaptic density protein isolation

Postsynaptic density (PSD) proteins were isolated as described (*63*). Briefly, frozen pooled hippocampi from four mice were homogenized in 1 mL of lysis buffer (0.32 M sucrose in 4 mM HEPES [pH 7.4], 5 mM sodium fluoride, 1 mM sodium orthovanadate, 1 mM sodium pyrophosphate, 1 mM β-glycerophosphate, and 1× protease inhibitor cocktail [P8340, MilliporeSigma]) using a Dounce homogenizer. A 50 μL aliquot of the homogenate was reserved as the total protein fraction (“Total”) by mixing with 2× Laemmli buffer. The remaining homogenate was centrifuged at 900 × g for 10 min at 4°C, and the resulting supernatant was subsequently centrifuged at 10,000 × g for 15 min at 4°C. The resulting pellet was washed by resuspension in lysis buffer and centrifuged again. The pellet was then resuspended in 1.4 mL H₂O and homogenized by passing through a 27-gauge needle five times. Subsequently, 6 μL of 1 M HEPES was added to achieve a final concentration of 4 mM, and the lysate was rotated for 30 min at 4°C. A 75 μL aliquot of the lysate was reserved as the crude synaptosome fraction (“Crude”) by mixing with 2× Laemmli buffer. The remaining lysate was centrifuged at 25,000 × g for 20 min at 4°C using a TLA100.3 rotor in an Optima TLX ultracentrifuge (Beckman Coulter). The pellet was resuspended in lysis buffer and layered onto a discontinuous sucrose gradient (0.8 M, 1.0 M, and 1.2 M sucrose in 4 mM HEPES), followed by centrifugation at 150,000 × g for 2 h at 4°C using an SW55Ti swinging-bucket rotor in an Optima XPN-100 ultracentrifuge (Beckman Coulter). The 700 μL interphase between the 1.0 M and 1.2 M sucrose layers, containing synaptic plasma membranes, was collected and diluted with 800 μL of 4 mM HEPES, followed by centrifugation at 150,000 × g for 30 min at 4°C using a TLA100.3 rotor. The resulting pellet was resuspended in 50 mM HEPES / 2 mM EDTA buffer, followed by the addition of 1.25 mL of 0.54% Triton X-100 in 50 mM HEPES / 2 mM EDTA buffer. After rotation for 15 min at 4°C, samples were centrifuged at 32,000 × g for 20 min at 4°C using a TLA100.3 rotor. The final pellet was collected as the postsynaptic density fraction (“PSD”) by mixing with 2× Laemmli buffer.

### Electrophysiology

Ten to twelve weeks age of WT control mice, global *Becn1*^+/-^ KO mice, conditional CamKII^Becn1^-KO mice, and CA3^Becn1^-KO mice were anesthetized by isoflurane and transcranially perfused with a modified ice-cold sucrose-based cutting solution (pH 7.3) containing 10 mM NaCl, 25 mM NaHCO3, 195 mM Sucrose, 5 mM Glucose, 2.5 mM KCl, 1.25 mM NaH2PO4, 2 mM Na-Pyruvate, 0.5 mM CaCl2, and 7 mM MgCl2, bubbled continuously with 95% O2 and 5% CO2 (*64*). The mice were then decapitated, and the entire brain was removed and immediately submerged in the cutting solution. The slices (250 µm) were cut with a Micron Leica VT1000s vibratome (Leica, IL, U.S.). Brain slices containing the hippocampal brain regions were obtained for each animal (Bregma -2.06 mm to -1.46 mm; Interaural 1.74 mm to 2.34 mm). The slices were recovered for 1 h at 34°C and then maintained at room temperature in artificial cerebrospinal fluid (aCSF, pH 7.3) containing 126 mM NaCl, 2.5 mM KCl, 2.4 mM CaCl2, 1.2 mM NaH2PO4, 1.2 mM MgCl2, 5.0 mM glucose, and 21.4 mM NaHCO3) saturated with 95% O2 and 5% CO2 before recording (*65*).

The brain slices were transferred to a recording chamber and allowed to equilibrate for at least 10 min before recording. The slices were perfused at 34°C in oxygenated aCSF at a flow rate of 1.8-2 ml/min. Hippocampal neurons were visualized using epifluorescence and IR-DIC imaging on an upright microscope (Eclipse FN-1, Nikon) equipped with a movable stage (MP-285, Sutter Instrument). For the miniature excitatory postsynaptic current (mEPSC) recordings, voltage-clamp recordings in the presence of low Mg2+ (MgCl2 in aCSF was reduced from 1.2 mM to 0.1 mM) to avoid Mg2+-block of NMDARs (*66*). The pipette solution contained 125 mM CsCH3SO3, 10 mM CsCl, 5 mM NaCl, 2 mM MgCl2, 1 mM EGTA, 10 mM HEPES, 5 mM (Mg)ATP, and 0.3 mM (Na)2GTP (pH 7.3 with NaOH). The mEPSCs were recorded in whole-cell voltage-clamp mode, by holding the membrane potential at Vh = − 60 mV in the presence of 1 μM TTX, 50 μM bicuculline (*65*). A baseline of average mEPSC total current amplitude was recorded and analyzed. NMDAR mEPSC amplitude was calculated by subtracting the average mEPSC total current amplitude in the presence of 50 μM D-AP5 from that recorded in its absence. AMPAR mEPSC amplitude was then calculated by subtracting the background current amplitude (recorded in the presence of 50 μM D-AP5 and 30 μM CNQX) from that recorded in the presence of D-AP5 only.

### Golgi-Cox staining

Golgi-Cox staining was performed using the FD Rapid GolgiStain Kit (FD NeuroTechnologies, #PK401) according to the manufacturer’s instructions. Bright-field images were acquired using a Nikon ECLIPSE Ti microscope (Nikon).

### Tandem affinity purification and proteomics of the BECN1 and BECN2 complexes

Tandem affinity purification was performed as described (*10*). Briefly, HEK293 cells stably expressing tetracycline-inducible ZZ-tagged hBECN1–3×FLAG and ZZ-tagged hBECN2–3×FLAG were generated using the pZZ-3×FLAG construct (*67*) and the Flp-In T-REx 293 cell system (R78007, ThermoFisher) according to the manufacturer’s protocol. Cells were cultured in medium containing 1 μg/mL tetracycline for 24 hours. After washing with PBS, cells were lysed in Triton X-100 lysis buffer. Lysates were centrifuged at 15,000 × g for 10 min at 4°C, and the supernatants were incubated with IgG Sepharose (17096901, Cytiva) for 3 hours at 4°C. Bound proteins were eluted by TEV protease cleavage (V6101, Promega). The eluate was then incubated with anti-FLAG affinity gel (A2220, MilliporeSigma) for 14-16 hours at 4°C, and bound proteins were eluted in Laemmli buffer. Immunoprecipitated proteins were identified by proteomic analysis performed at the Northwestern University Proteomics Core Facility and Creative Proteomics.

### Plasmids

Double-stranded oligonucleotides encoding shRNAs targeting specific genes were cloned into the AgeI and EcoRI restriction sites of the pLKO.1-TRC plasmid (Addgene plasmid #10878). The gene-specific shRNA sequence corresponding to human *BECN1* was CTCAAGTTCATGCTGACGAAT. A scramble shRNA in pLKO.1 (Addgene plasmid #1864) was used as a negative control.

For CRISPR/Cas9 knockout experiments, double-stranded oligonucleotides encoding guide RNAs (gRNAs) targeting specific genes were cloned into the BbsI sites of the pSpCas9(BB)-2A-Puro (PX459) V2.0 plasmid (Addgene plasmid #62988). The gene-specific gRNA sequence corresponding to human *BECN1* was GGACACGAGTTTCAAGATCC.

FLAG-BECN2, FLAG-BECN1 full-length (FL), and FLAG-BECN1 deletion mutant plasmids were described (*10, 68*). HA-BECN1 was generated by subcloning into the EcoRI site of the pcDNA3.1(−) plasmid, replacing the FLAG tag of FLAG-BECN1 with an HA tag using the In-Fusion cloning kit (Takara Bio, #6389110). The 3×FLAG-UBE3A construct was generated by replacing the HA tag of pLVX-HA-UBE3A-IRES-Puro (Addgene #107349) with a 3×FLAG tag and subcloning into the EcoRI site of the pcDNA3.1(−) plasmid using the In-Fusion cloning kit. The Myc-Arc plasmid was generated by cloning codon-optimized synthetic DNA (GENEWIZ) into the EcoRI site of the pcDNA3.1(−) plasmid using the In-Fusion cloning kit. The codon-optimized Myc-Arc sequence is shown below.

Human RNF40 and DZIP3 were cloned from cDNA libraries generated by reverse transcription of total RNA isolated from WI-38 cells. FLAG-tagged RNF40 (FLAG-RNF40) and DZIP3 (FLAG-DZIP3) were subcloned into the EcoRI restriction site of the pcDNA3.1(−) vector using the In-Fusion cloning kit. pcDNA-gp78-FLAG (Addgene #62370), pSMPP-mCherry-hTRIM21 (Addgene #104972), and pCMV3TAG8li-hRNF160-3FLAG (Addgene #159138) were obtained from Addgene.

MYC-ARC/Arg3.1:

atggaacaaaaacttattagcgaagaagatcttgaactggat catagaacatctggaggtttgcatgcatatcctggtccaagaggtg gacaggtagccaaacctaatgtgattctccagattggaaaatg tagagctgaaatgcttgaacatgttagaaggactcatagacatttg ttggcagaagtgtccaaacaagttgaaagagaattgaa gggactgcatagatctgtgggtaaattggaaagtaatcttgacggatacg ttccaactagcgattcacaaaggtggaagaagtccattaaggcctgcctttgcag atgtcaggaaacaatcgccaatctggaaagat gggtcaaaagggagatgcacgtgtggagggaagttttctacagattgga aagatgggcagatagacttgaatctacaggaggcaaa tatcctgttggttctgaatcagctagacacacagtaagtgtgggtgtaggaggtccagaatcttattgtcatgaagccgatgggtatgatt atacagtgtctccatacgcaattacacctcctccagctg ctggagaacttccaggtcaggaaccagcggaagcccagcagtatcagc catgggtgcctggtgaggatgggcaaccttcacctggtg ttgatacacagatttttgaagatcccagagaatttctgtctcatcttgaaga atatcttcggcaggtaggaggttctgaagaatattggctgtctcaaatacaaaa tcatatgaatggaccagctaaaaagtggtgggaat tcaagcaaggaagtgtaaaaaactgggtggagtttaaa aaagaatttcttcaatattctgaaggaacactttctagagaagctatacaa agagaactcgatctgccacaaaaacagggagaaccattggaccaatttctgtg gaggaagagagatctttatcaaactctttatgtgg atgcagatgaggaagaaattatccagtatgttgtcggaactctcca accaaaactcaagagatttttgagacatccactgcctaaaac actcgaacaattgatccaaagaggaatggaa gtccaggatgaccttgaacaagcggctgaacctgctggacctcatttgccagtaga agatgaagctgaaacactgacacctgcacctaattctgaaagtgtagcaagtgatagaacacaaccggagtag

### Generation of human *BECN1* constructs

For in vitro experiments, HA-tagged human BECN1 C375R and F372S mutant expression plasmids were generated by site-directed mutagenesis from the pcDNA3.1(−) HA-hBECN1 wild-type plasmid using the In-Fusion cloning kit. For human *BECN1* mRNA rescue experiments in zebrafish, Gateway-compatible full-length WT *BECN1* Open Reading Frame (ORF) (Dharmacon; clone # OHS5893-202492806; GenBank: NM_001313998.2) was transferred to a Gateway compatible pCS2+ destination vector by LR-clonase II mediated recombination (Thermo Fisher Scientific). Site-directed mutagenesis was performed to generate plasmids harboring *BECN1* variants. WT and mutant constructs were linearized with *Not*I and *in vitro* transcribed into mRNA with the mMessage mMachine SP6 Transcription kit (Ambion), according to the manufacturer’s instructions.

### Cells

HEK293, HEK293T, and WI-38 cells were cultured in Dulbecco’s modified Eagle’s medium (DMEM) with 10% fetal bovine serum (FBS), 100 units/ml penicillin, and 100 μg/ml streptomycin. *BECN1* knockout cells were generated by transfecting CRISPR/Cas9 knockout plasmids, followed by puromycin selection. For protein transient expression, plasmids were transfected using Lipofectamine 3000 (L3000, Thermo Fisher Scientific). In some experiments, cells were treated with 10 μM MG132 (F1101, UBPBio) for the indicated time before sample collection.

### Lentivirus production and infection

HEK293T cells were transfected with a lentiviral vector together with pCMV-VSV-G (Addgene plasmid #8454) and psPAX2 (Addgene plasmid #12260) using Lipofectamine 3000 (L3000, Thermo Fisher Scientific). After 14–16 hours, the medium was replaced with fresh medium. Forty-eight hours post-transfection, the medium containing lentivirus was collected and filtered. Target cells were infected by incubation in lentivirus-containing medium with 8 μg/ml polybrene, followed by selection with appropriate antibiotics.

### Immunoblotting of cell lines and mouse tissues

Mouse tissues or PBS-washed cultured cells were lysed in RIPA buffer (20 mM Tris-HCl [pH 7.4], 150 mM NaCl, 1% NP-40, 0.1% SDS, 0.5% sodium deoxycholate, 1 mM EDTA, 5 mM sodium fluoride, 1 mM sodium orthovanadate, 1 mM sodium pyrophosphate, 1 mM β-glycerophosphate, 1× protease inhibitor cocktail [P8340, MilliporeSigma]). Lysates were centrifuged at 15,000 × g for 10 min at 4°C, and supernatants were collected. Protein concentrations were determined using a BCA assay kit (23225, Thermo Fisher Scientific). Proteins were denatured in Laemmli buffer by heating at 95°C for 5 min. Equivalent amounts of protein were resolved by SDS-PAGE and transferred to PVDF or nitrocellulose membranes. Membranes were blocked with 0.5% skim milk or 5% BSA in TBS-T (Tris-buffered saline with 0.05% Tween-20) and probed with primary antibodies overnight at 4°C. After washing three times for 10 min each with TBS-T, membranes were incubated with HRP-conjugated secondary antibodies for 2 hours at 4°C. Specific bands were visualized by chemiluminescence using the ChemiDoc MP Imaging System (Bio-Rad), and band intensities were quantified using ImageJ.

### Immunoblotting of *becn1* zebrafish mutants

Heterozygous *becn1* adults (F1s) were in-crossed and euthanized at 8 dpf. Larvae were decapitated with microscissors and we collected individual tails for genotyping and individual heads for protein extraction. Genotype-matched WT and homozygous mutant *becn1* heads were pooled (25 heads/condition) in RIPA lysis buffer (50 mM HEPES [pH 7.6], 1% Triton X-100, 0.1% SDS, 50 mM NaCl, 0.5% sodium deoxycholate, 1 mM NaF, and 0.02 mM DTT) (Sigma Aldrich). The heads were ground with a sterile pestle in 1.5ml tubes and centrifuged for 3 minutes at 4C. Total protein lysates were isolated, quantified by BCA assay (Thermo Fisher Scientific) and 50 μg of heat denatured protein lysate mixed with Laemmli sample buffer was migrated on a 4–15% mini-PROTEAN TGX Precast Gel (Bio-Rad). After separation, proteins were transferred to a polyvinylidene difluoride (PVDF) membrane and incubated overnight in primary rabbit antibody solution in 3% BSA in PBS (Becn1, NSJ Bioreagents, cat#F53282-0.4ML; 1:1000) and actin was used as a loading control (Santa Cruz Biotechnology, cat#sc-1615, 1:3000) followed by incubation in secondary antibody solution containing 3% BSA in PBS (Becn1, anti-mouse IgGκ-HRP, Santa Cruz Biotechnology, sc-516102, 1:5,000 dilution; actin, anti-goat IgG-HRP, cat #sc-2354, 1:4,000). We detected immunoblots using Super Signal West Pico PLUS chemiluminescent substrate (Thermo Fisher Scientific) according to manufacturer’s instructions and chemiluminescent images were obtained on a ChemiDoc Imaging System (Bio-Rad).

### Immunoprecipitation and ubiquitinated proteins pull down

Mouse tissues were lysed in Triton X-100 lysis buffer (1% Triton X-100, 20 mM HEPES [pH 7.4], 150 mM NaCl, 1 mM EDTA, 5 mM sodium fluoride, 1 mM sodium orthovanadate, 1 mM sodium pyrophosphate, 1 mM β-glycerophosphate, and 1×protease inhibitor cocktail [P8340, MilliporeSigma]). Lysates were centrifuged at 15,000 × g for 10 min at 4°C, and the supernatants were pre-cleared by incubation with 3 μg of mouse IgG isotype control (10400C, Thermo Fisher Scientific) and Protein A/G beads (sc-2003, Santa Cruz Biotechnology) for 1 h at 4°C. The pre-cleared lysates were incubated with 2 μg of anti-BECN1 antibody (sc-48341, Santa Cruz Biotechnology) or mouse IgG control, together with Protein A/G beads, for 14–16 h at 4°C. For total ubiquitinated protein pull-down, the pre-cleared lysates were incubated with agarose-coupled Tandem Ubiquitin Binding Entity (TUBE2) resin (UM402, LifeSensors) for 14–16 h at 4°C. Beads were washed five times with lysis buffer, and immunoprecipitated or pull-down proteins were eluted in Laemmli buffer for immunoblotting.

HEK293 cells transfected with plasmids using Lipofectamine 3000 were lysed in Triton X-100 lysis buffer. Lysates were centrifuged at 15,000 × g for 10 min at 4°C and incubated with anti-HA-tag (sc-7392) antibody and Protein A/G beads (sc-2003, Santa Cruz Biotechnology) or anti-Flag M2 affinity gel (A2220, Millipore Sigma), or anti-Bcl2 agarose beads (sc-7382 AC) for 14–16 h at 4°C. Beads were washed five times with lysis buffer, and immunoprecipitated proteins were eluted in Laemmli buffer and analyzed by immunoblotting.

### Immunofluorescence staining

Mouse brains were harvested from mice perfused with 4% paraformaldehyde (PFA) in PBS and fixed in 4% PFA/PBS for 4 hours. Tissues were incubated sequentially in 15% and 30% sucrose/PBS overnight for cryoprotection, embedded in optimal cutting temperature (OCT) compound, and sectioned at 40 μm using a cryomicrotome (CM1860, Leica Biosystems). Sections were washed twice with PBS for 5 min, followed by blocking in 5% normal goat serum (NGS) with 0.3% Triton X-100 in PBS for 60 min at room temperature. Sections were incubated with primary antibodies in antibody buffer (1% BSA, 0.3% Triton X-100 in PBS) overnight at 4°C, washed three times with PBS buffer, and incubated with Alexa Fluor-conjugated secondary antibodies for 2 hours at room temperature. Sections were mounted with ProLong Diamond Antifade mountant (P36961, Thermo Fisher Scientific). Fluorescence images were acquired using a spinning-disk confocal microscope (X-Light V2 spinning disk confocal system [CrestOptics] with Hamamatsu Flash 4 camera [Hamamatsu Photonics] mounted to a Nikon ECLIPSE Ti microscope [Nikon]).

### Quantitative PCR (qPCR)

Total RNA was isolated from mouse hippocampus using TRIzol Reagent (15596018, Thermo Fisher Scientific), and 2 μg of total RNA was reverse-transcribed using the High-Capacity cDNA Reverse Transcription Kit (4368814, Thermo Fisher Scientific) according to the manufacturer’s instructions. qPCR was performed using PowerUP SYBR Green Master Mix on a QuantStudio 6 Pro Real-Time PCR System (Thermo Fisher Scientific). Relative gene expression was calculated using the 2^-ΔΔCt^ method. The following gene-specific primers were used; mouse *GluA1*-5’ ATTGACATATCCCGGCGAGG; mouse *GluA1*-3’ TTCCAGTAACCAATCTTGCGGA; mouse *GluA2*-5’ CAAATGGACCCCGGAAGATTG; mouse *GluA2*-3’ ACAGTAGCCCTCATAACGCTC; mouse *Arg/Arg3.1*-5’ CTGAAGCAGCAGACCTGACA; mouse *Arg/Arg3.1*-3’ CAGCAGCCTTGAGACCTGG; mouse *Rplp0*-5’ TTCGTGTTCACCAAGGAGGAC; mouse *Rplp0*-3’ ATGATCAGCCCGAAGGAGAAG.

### Antibodies to human and mouse proteins

The following antibodies were used in immunoblotting: anti-Flag M2 antibody (F1804) (Millipore Sigma); anti-Synaptophysin (ab8049) antibody (Abcam); anti-p62 (610833) antibody (BD Biosciences); anti-LC3B (NB100-2220) antibody (Novus); anti-BECN1 (11306-1-AP), and anti-HUWE1 (19430-1-AP) antibodies (Proteintech); anti-Arc (sc-17839), anti-Myc (sc-40), anti-UBE3A (E6-AP) (sc-25509), anti-GAPDH (sc-365062), anti-VPS34 (sc-365404), anti-Ubiquitin (sc-8017), HRP-conjugated anti-Bcl2 (sc-7382 HRP), and HRP-conjugated anti-β-actin (sc-4778 HRP) antibodies (Santa Cruz Biotechnology); anti-AMBRA1 (24907), anti-AMPAR1 (GluA1) (13185), anti-AMPAR2 (GluA2) (5306), anti-CREB (9197), anti-Phospho-CREB (Ser133) (9198), anti-HA-tag (3724), anti-PSD95 (3450), anti-NMDAR1 (GluN1) (5704), anti-UVRAG (5320), and HRP-conjugated anti-Mouse IgG light-chain specific (91196) antibodies (Cell Signaling Technology); HRP-conjugated anti-Rabbit IgG (111-035-003), HRP-conjugated anti-Mouse IgG (115-035-003), and HRP-conjugated anti-Rabbit IgG light chain specific (211-032-171) antibodies (Jackson ImmunoResearch).

### Sequence alignment and 3D structure prediction

Sequence alignment of BECN1 orthologs was performed using the Clustal Omega method in SnapGene software. Three-dimensional protein structures of BECN1 WT, C375R, and F372S were predicted using AlphaFold3 (https://alphafoldserver.com) (*69*). The predicted structures were aligned in three dimensions using Pairwise Structure Alignment (https://www.rcsb.org/alignment) (*70*).

### Quantification and statistical analysis

All data are presented as mean ± standard error of the mean (SEM). Statistical comparisons between two groups were performed using Student’s t-test. For comparisons involving more than two groups, two-way analysis of variance (ANOVA) followed by Bonferroni’s or Dunnett’s post hoc test were used. All zebrafish morphometric data were analyzed with an unpaired student’s t-test. Experiments were performed at least twice with the investigator masked to experimental conditions. Analyses were conducted using GraphPad Prism (Dotmatics). Differences with a p-value < 0.05 were considered statistically significant.

## Data Availability

All data produced in the present study are available upon reasonable request to the authors

## Acknowledgements

We are grateful to the participating families for their enthusiasm and interest in our research. We thank the following facilities at Northwestern University Feinberg School of Medicine for technical support: the Proteomics Core for proteomics studies, the Behavioral Phenotyping Core for mouse behavioral analyses, and the NUSeq Core of Center for Genetic Medicine for sequencing. E.E.D. is the Ann Marie and Francis Klocke, MD Research Scholar. The work was supported by US National Institutes of Health grants R01 DK129548, R56 DK133776 and R01 DK142603 (Y.H.), R01 MH106826 (E.E.D.), R01 DK113170, R01 DK123447 and R01 DA056720 (C.H.).

## Author Contributions

Project conceptualization: E.E.D., C.H. Experimental performance: K.K., F.U., P.L., I.M.R., B.F., M.S., M-S.K., H.D., M.J.O, Z.Q, A.B., Y.H. Data analysis: K.K., F.U., P.L., I.M.R., B.F., M.S., M-S.K., Y.H., C.H. Clinical ascertainment of human data: L.A., M.A.C., A.S., A. Mercier, A. McRae, O.A., M.O.H, A.A., S.M., F.S.A. Writing – original draft: K.K., F.U., E.E.D., C.H. Writing – review & editing: L.A., A.C., F.S.A., Y.H., E.E.D., C.H.

